# Predictors of mortality, ICU admission and intubation among hospitalized COVD-19 patients in Jordan

**DOI:** 10.1101/2022.01.31.22270218

**Authors:** Khaled Al Oweidat, Asma S. Albtoosh, Ahmad A. Toubasi, Mona Khaled Ribie, Manar M. Hasuneh, Daniah L. Alfaqheri, Abdullah H. Alshurafa, Mohammad Ribie, Nathir Obeidat

**Affiliations:** Department of Respiratory and Sleep Medicine, Department of Internal Medicine, School of Medicine, The University of Jordan, Amman, Jordan; Faculty of Medicine, The University of Jordan, Amman, Jordan; Department of Internal Medicine, School of Medicine, The University of Jordan, Amman, Jordan

## Abstract

**Introduction:** Several studies characterized COVID-19 patients and revealed the factors associated with COVID-19 mortality and severity. However, few studies were conducted in the Middle East, hence we decided to conduct this study to characterize COVID-19 patients, identify the factors associated with COVID-19 mortality and severity and to compare the patients’ outcomes in the two waves of the pandemic.

**Method:** We conducted a retrospective single center cohort study among COVID-19 patients admitted to our hospital. The primary outcomes were COVID-19 related death, ICU admission and invasive intubation. Whereas, the secondary outcome was the length of hospital stay. In addition, the patients were divided into two cohorts to represent the two waves of the pandemic, to compare between them based on the outcomes.

**Results:** The total number of COVID-19 patients included in our study was 745. The overall rates of COVID-19 related mortality, ICU admission and invasive intubation were 23.0%, 28.3% and 10.8%, respectively. CKD, troponin, LDH and O2 saturation <90% were significant predictors for mortality. Moreover, the significant predictors for ICU admission were HF and remdesivir use. Whereas, O2 saturation<90% and GI symptoms were the only significant predictors for invasive intubation. Additionally, the second wave had significantly higher rates of ICU admission compared to the first one.

**Conclusion:** Our study showed that several factors predicted COVID-19 mortality, ICU admission and invasive intubation. Additionally, our study revealed that the second wave of the pandemic had significantly higher rates of ICU admission but not mortality or invasive intubation compared to the first wave.

## Introduction

A novel coronavirus (CoV) named severe acute respiratory syndrome coronavirus 2 (SARS-CoV-2) by the World Health Organization (WHO) is responsible for the recent pneumonia pandemic that started in early December, 2019 in Wuhan City, Hubei Province, China [1, 2]. The transmission of SARS-CoV-2 occurs mainly through respiratory droplets and aerosols and causes a disease that is now referred to as COVID-19 [3]. The disease was classified based on the diverse clinical features it can present with into asymptomatic, mild, moderate, severe and critical. Asymptomatic infection was defined as having no symptoms that are consistent with COVID-19. When individuals presented with the signs and symptoms of COVID-19 without shortness of breath or abnormal chest imaging, the illness was classified as mild. Moderate illness was defined as individuals who show evidence of lower respiratory disease during clinical assessment or imaging and who have an oxygen saturation (SpO_2_) ≥94% on room air at sea level. Severe illness was defined as individuals who have SpO_2_ <94% on room air at sea level, a ratio of arterial partial pressure of oxygen to fraction of inspired oxygen (PaO_2_/FiO_2_) <300 mm Hg, a respiratory rate >30 breaths/min, or lung infiltrates >50%. Lastly, the definition of critical illness was made in individuals who have respiratory failure, septic shock, and/or multiple organ dysfunction [4].

As of January 2022, over 350 million COVID-19 cases have been reported worldwide resulting in upwards of 5 million deaths [5]. Mortality rate early in the pandemic was high among hospitalized patients approaching 32% [6]. Published research on COVID-19 disease described many factors that predict severe and fatal COVID-19 disease, including: demographic factors as well as clinical, immunological, hematological, biochemical and radiographic findings [7]. Demographic factors such as age and gender have been reported to impact COVID-19 outcomes with older age predicting disease severity and survival [7–11] and male sex associated with a severe clinical course and mortality [8]. Preexisting comorbid conditions such as cardiovascular disease, diabetes, chronic respiratory disease, hypertension, and cancer were also associated with increased case fatality rate [9–15]. Moreover, high levels of C-reactive proteins (CRP), D-dimer, procalcitonin (PCT), total bilirubin, renal dysfunction indices and interleukin-6 level (IL-6) as well as low lymphocyte count were found to predict severe disease course and death [12]. On the other hand, patients who were treated with methylprednisolone had lower mortality rates than patients who were not treated with the drug [12].

Until the 28^th^ of January 2022, Jordan had 13,157 COVID-19 deaths and a total of 1,189,080 cases [5]. In the Middle East, a few studies revealed the clinical characteristics [16] and predictors of COVID-19 severity and mortality [17]. To our best knowledge, no studies were conducted in Jordan to reveal the predictors of COVID-19 mortality and severity. Accordingly, the scarcity of data about COVID-19 in the Middle East necessitates the conduct of studies that characterize COVID-19 patients in the region. The aim of this study was to describe the clinical characteristics of COVID-19 in hospitalized patients and identify risk factors associated with mortality, ICU admission, intubation rate and length of stay in those patients. In addition, our study aimed to compare the outcomes between the two admission waves of the pandemic in our hospital.

## Material and Methods

### Study population

A trained team of physicians and medical students performed a retrospective study among hospitalized COVID-19 patients at Jordan University Hospital (JUH) in Amman, Jordan using the hospital’s electronic medical records (EMR). Data was extracted from a total of 753 medical records of COVID-19 patients with a laboratory confirmed diagnosis of SARS-CoV-2 infection by nasopharyngeal swab real-time reverse-transcriptase polymerase chain reaction (RT-PCR). The RT-PCR was performed using Zybio nucleic acid extraction test, China and the detection kit was Zybio nucleic acid detection kit. The RT-PCR samples were collected within the first two weeks of the symptoms’ onset. All patients who were admitted to JUH between September 2020 to August 2021 were eligible for inclusion in the study. The patients were divided into two cohorts to represent the two waves of the pandemic in Jordan and, to subsequently compare between them based on the outcome measures. The first wave of the pandemic in Jordan was from September, 2020 to January, 2021 while the second wave was between February, 2021 and August, 2021.

### Data Collection

The following patients’ data was collected:

- Patient’s characteristics, including: demographics (age, sex and smoking status), presenting symptoms (cough, shortness of breath, runny nose, fever, headache and gastrointestinal symptoms) and comorbidities (hypertension (HTN), diabetes mellitus (DM), cardiovascular disease (CVD), heart failure (HF), chronic kidney disease (CKD), malignancy, asthma, chronic obstructive pulmonary disease (COPD), dyslipidemia, neurological diseases and autoimmune diseases). In addition, patients’ presentation with fever or hypoxia was documented. Presence of fever on admission was identified by an emergency room triage oral temperature higher than 37.7°C [18], with hypoxia on admission determined by oxygen saturation and subsequently divided into 3 groups; <90%, 90%-94%, and >94% on room air or ambient air.
- Patient’s venous blood sample laboratory results were collected. These included complete blood count (CBC), D-dimer, C-reactive protein (CRP), Interleukin 6 (IL6), Troponin I, Procalcitonin, absolute lymphocyte count, ferritin level, serum creatinine, urea, last hemoglobin A1c (HbA1C) within 3 months of admission. In addition, lactate dehydrogenase (LDH), aspartate aminotransferase (AST), alanine aminotransferase (ALT), albumin, serum potassium, and serum sodium levels were also collected. Serum creatinine values were used to identify Acute kidney injury (AKI) among COVID-19 patients which was defined as an increase in serum creatinine to ≥1.5 times the baseline [19]. Moreover, troponin values were considered positive if they were ≥45 pg/ml. Furthermore, the HbA1C within the last 3 months of the admission was used to determine the state of diabetes control among patients with diabetes, with an HbA1C ≥9 indicating uncontrolled diabetes.
- Radiological features, including chest x-ray, high-resolution computed tomography (HRCT) and computed tomography pulmonary angiogram (CTPA).
- In-hospital management details, including medication received during admission (dexamethasone, antiviral therapy, tocilizumab) and oxygen delivery devices such as nasal cannula, face mask, venturi, non-rebreather mask, non-invasive ventilation, high flow nasal cannula and invasive mechanical ventilation.
- Patient’s outcomes during the entire hospitalization period including death, ICU admission, intubation and length of stay.

### Outcome Measures

The primary outcomes were COVID-19 related death, ICU admission and invasive intubation. Whereas, the secondary outcome was the length of hospital stay. COVID-19 related death was defined as mortality that occurred within the first 28 days from the admission and was attributed to one of the COVID-19 complications.

### Ethical Statement

Given the retrospective nature of the study, informed consent was not required. The study protocol was approved by the Institutional Review Board at JUH.

### Statistical Analysis

The patients’ data was entered on Microsoft Office Excel 2019, then imported into IBM SPSS v.25 software which was used to conduct the analysis. Counts and percentages were used to describe categorical variables whereas means and standard deviations were used to describe continuous variables. To identify the predictors of COVID-19 related mortality, ICU admission and invasive intubation, univariate binary logistic regression analysis was used. In addition, univariate linear regression was used to identify the predictors of length of hospital stay. To adjust for confounding variables, significant predictors of COVID-19 related mortality, ICU admission and invasive intubation were reexamined using multivariate binary logistic regression. While significant predictors of length of hospital stay were retested using multivariate linear regression. Results of univariate binary and linear logistic regression were expressed using crude odds ratio (COR) and crude B coefficient (CB) with their corresponding 95% confidence intervals (95% CIs), respectively. While the results of the multivariate binary and linear logistic regression models were expressed using adjusted odds ratio (AOR) and adjusted B coefficient (AB) with their corresponding 95% confidence intervals (95% CIs), respectively. All the variables with a P-value<0.05 in the univariate and multivariate logistic regression models were considered statistically significant.

## Results

### General Characteristics of the Included Patients

We retrieved 753 medical records of COVID-19 positive patients admitted in our hospital, of which 8 records were excluded due to missing data about their outcomes. The total number of the COVID-19 patients included in our study was 745 patients. The detailed characteristics of the patients are described in Table 1. The percentage of males in the study was 51.3% (382/745). The mean of the patients’ age was 63.15 ± 15.99 and the majority of the patients were non-smokers (86.6%). Moreover, the most frequent chief complaints reported by the patients were shortness of breath (35.0%), cough (15.5%) and generalized weakness (15.0%). In addition, 30.4% of the patients reported gastrointestinal symptoms (226/744). Regarding comorbidities, 44.8% of the patients had Diabetes Mellitus (DM) and 55.1% had Hypertension (HTN). Furthermore, 16.1% of the patients had coronary artery disease (CAD) and 8% of the patients had heart failure (HF). Additionally, 12.5% and 5.9% of the patients had chronic kidney disease (CKD) and neurologic diseases, respectively. Moreover, only 3.2% of the patients were vaccinated and the majority (64.4%) of the patients were from the second wave of the pandemic.

**Table 1.**
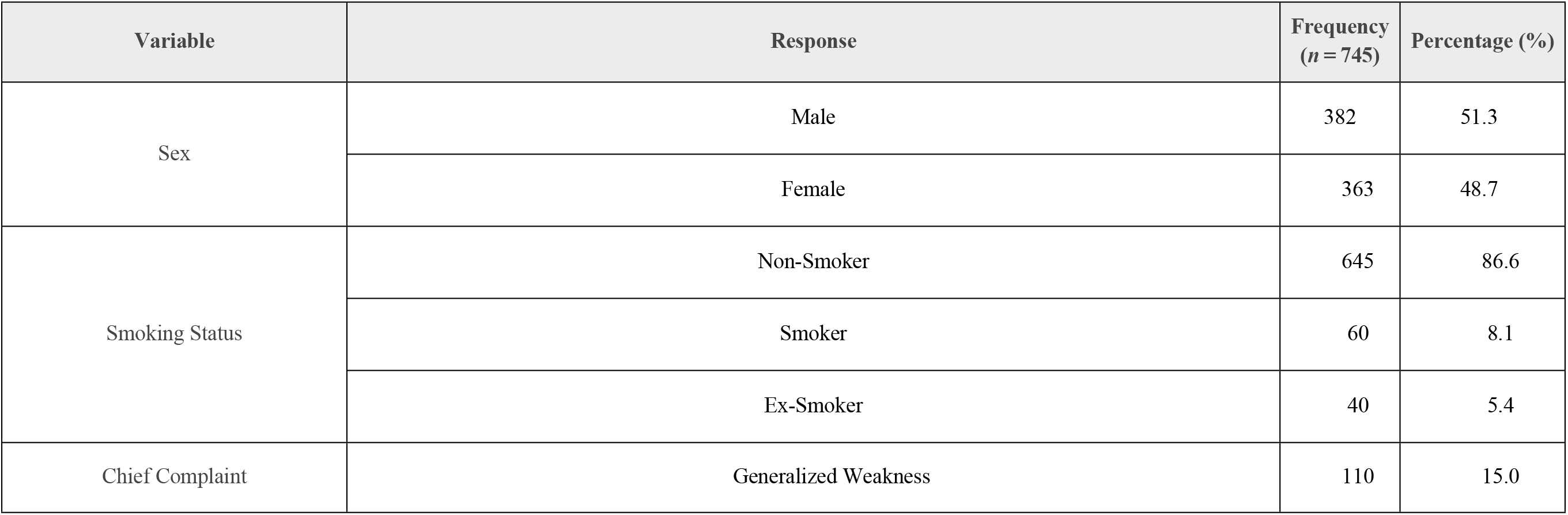
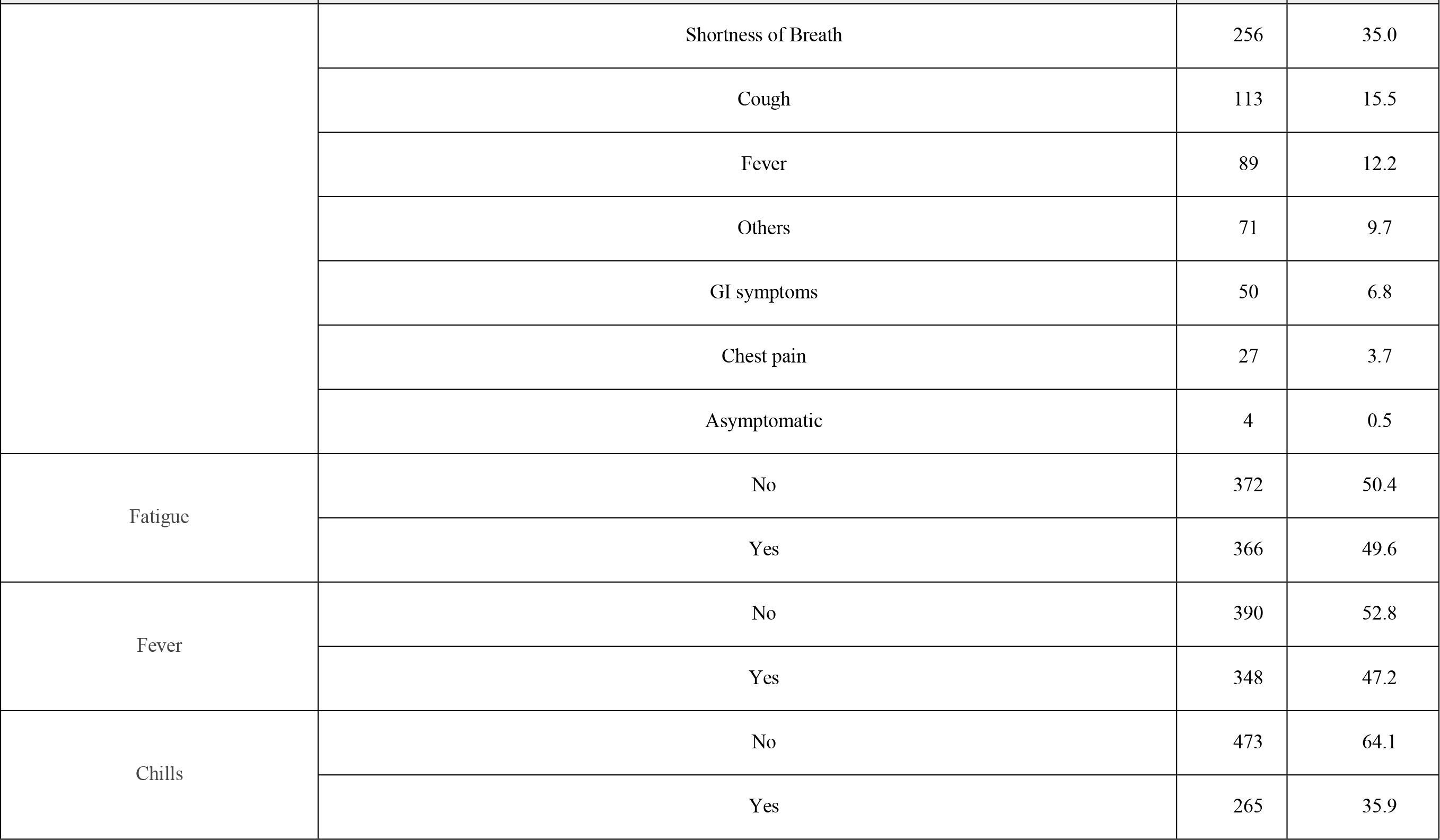
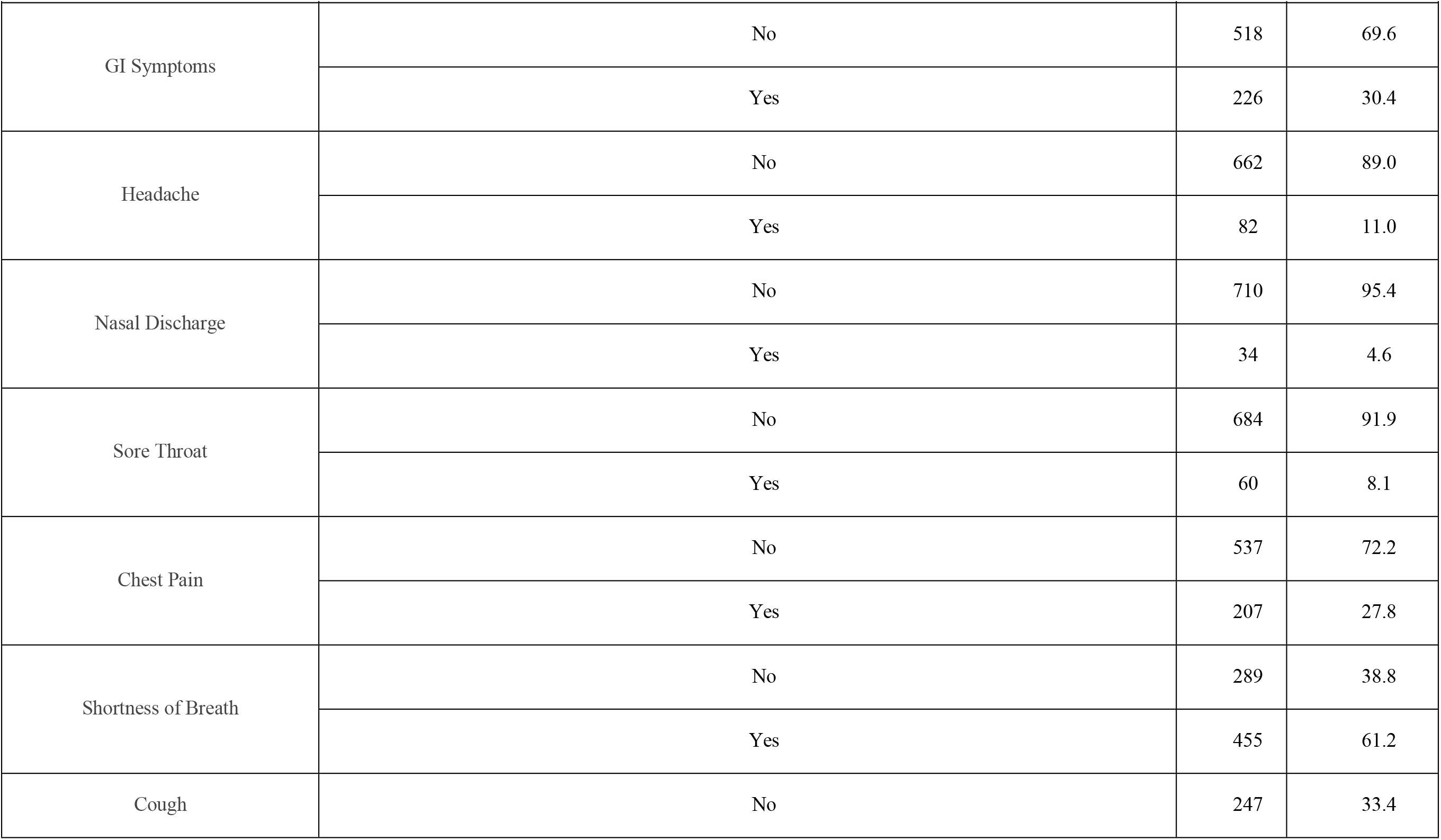
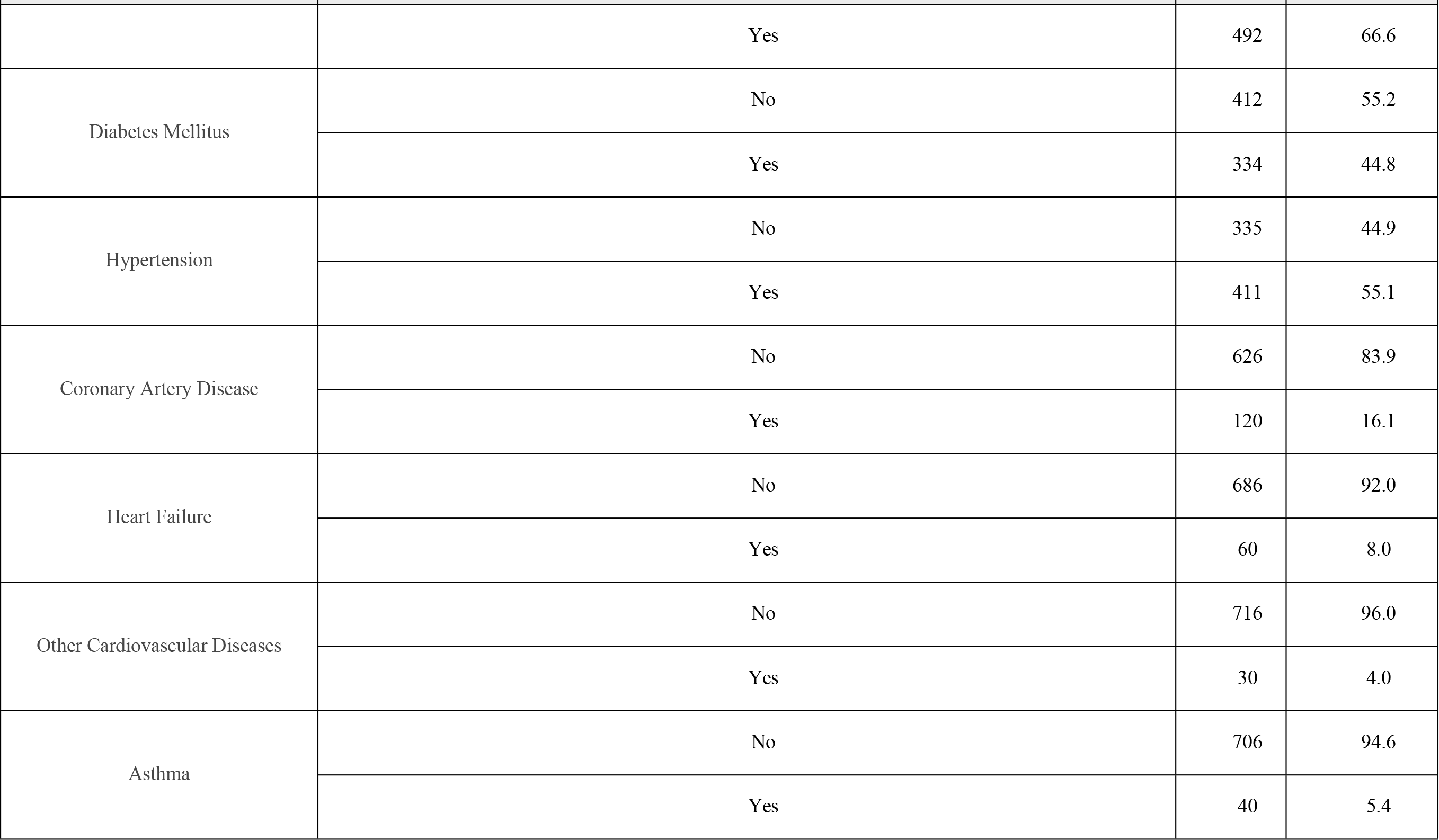
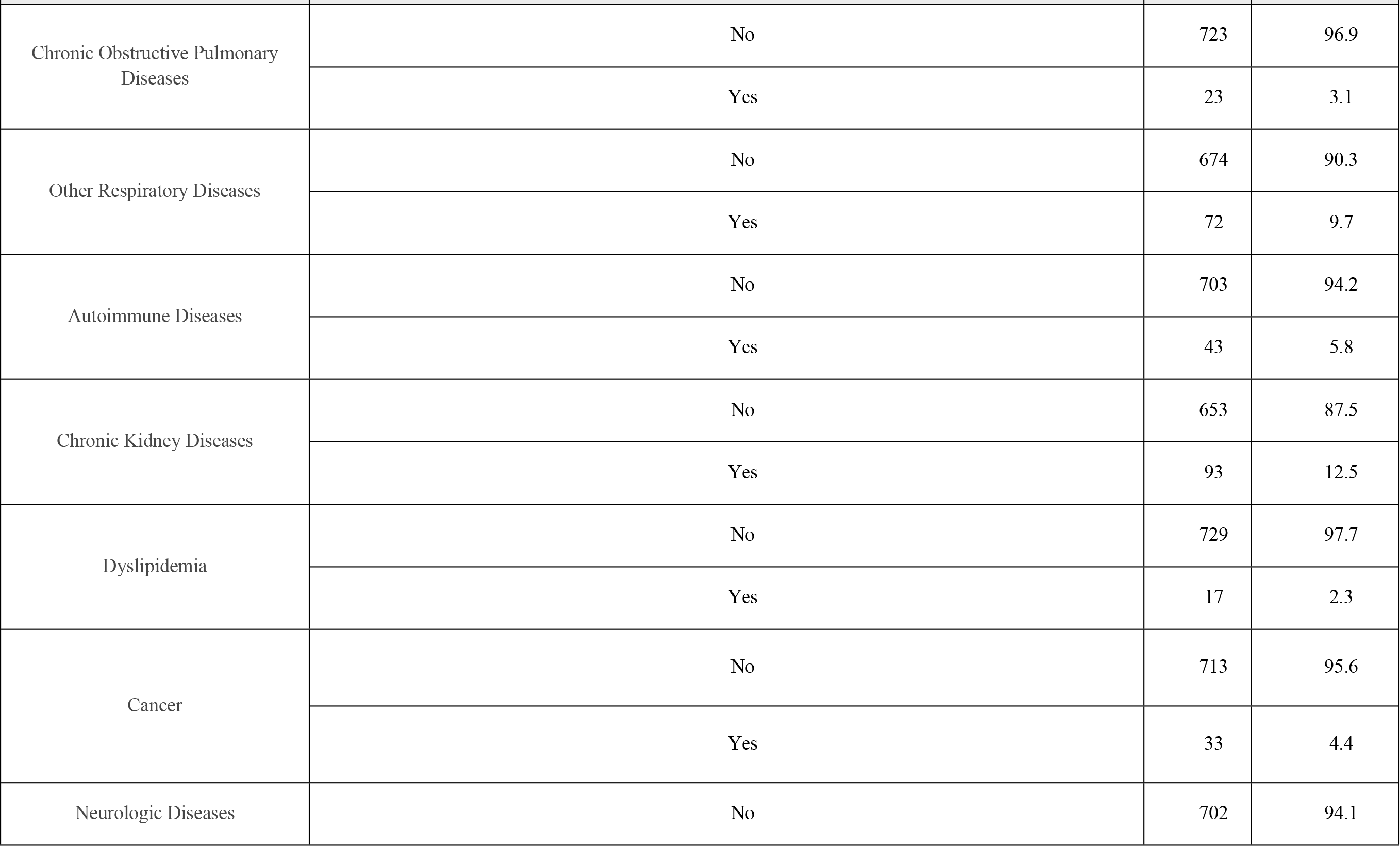
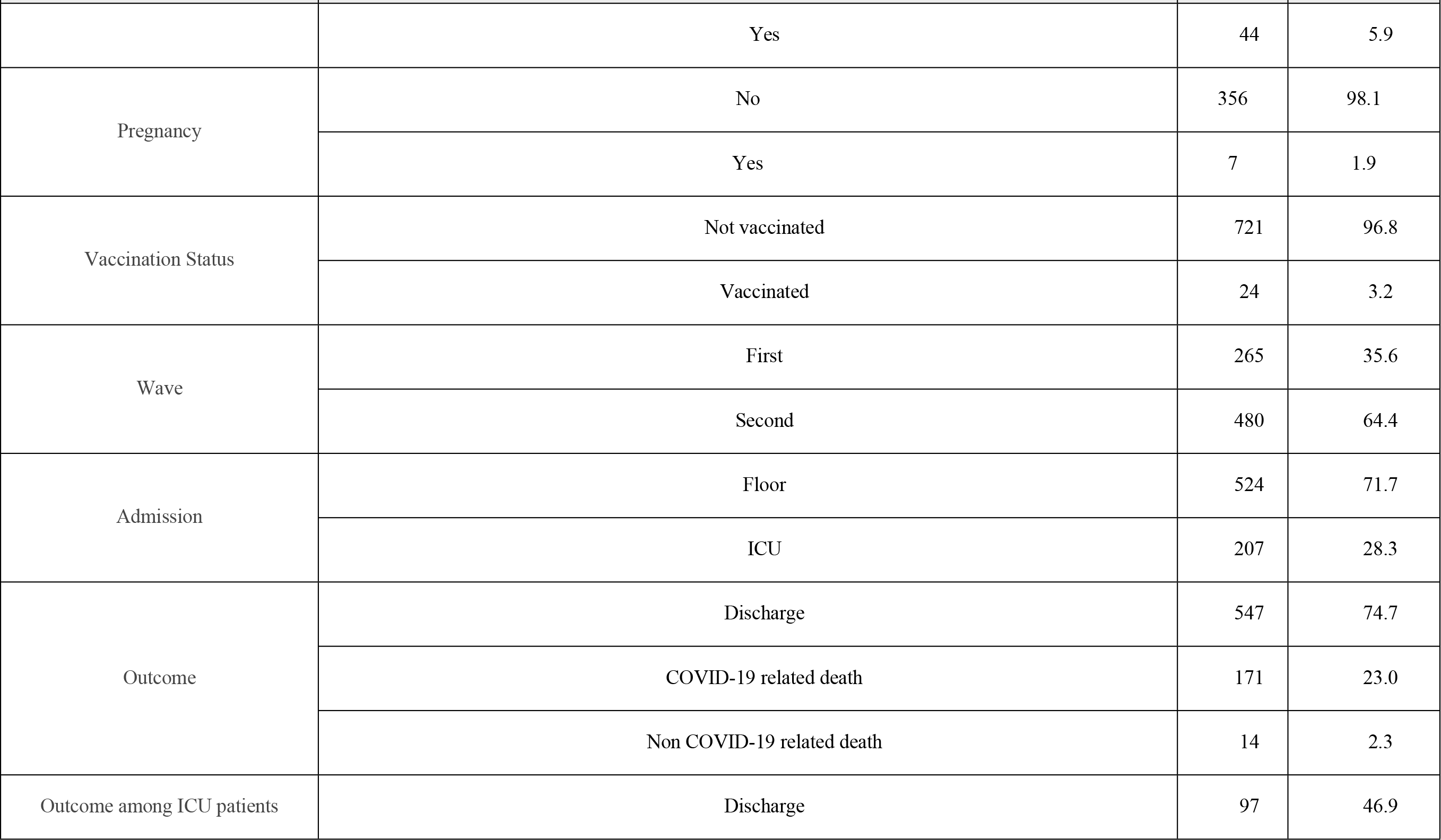
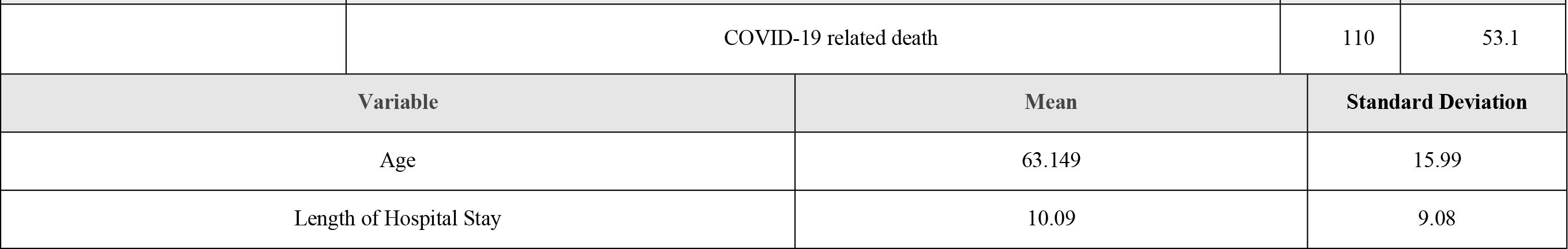
The General Demographics of the Patients.

| Variable | Response | Frequency<br>(n = 745) | Percentage (%) |
| --- | --- | --- | --- |
| Sex | Male | 382 | 51.3 |
|  | Female | 363 | 48.7 |
| Smoking Status | Non-Smoker | 645 | 86.6 |
|  | Smoker | 60 | 8.1 |
|  | Ex-Smoker | 40 | 5.4 |
| Chief Complaint | Generalized Weakness | 110 | 15.0 |

| Variable | Response | Frequency<br>(*n* = 745) | Percentage (%) |
| --- | --- | --- | --- |
|  | Shortness of Breath | 256 | 35.0 |
|  | Cough | 113 | 15.5 |
|  | Fever | 89 | 12.2 |
|  | Others | 71 | 9.7 |
|  | GI symptoms | 50 | 6.8 |
|  | Chest pain | 27 | 3.7 |
|  | Asymptomatic | 4 | 0.5 |
| Fatigue | No | 372 | 50.4 |
|  | Yes | 366 | 49.6 |
| Fever | No | 390 | 52.8 |
|  | Yes | 348 | 47.2 |
| Chills | No | 473 | 64.1 |
|  | Yes | 265 | 35.9 |
| GI Symptoms | No | 518 | 69.6 |
|  | Yes | 226 | 30.4 |
| Headache | No | 662 | 89.0 |
|  | Yes | 82 | 11.0 |
| Nasal Discharge | No | 710 | 95.4 |
|  | Yes | 34 | 4.6 |
| Sore Throat | No | 684 | 91.9 |
|  | Yes | 60 | 8.1 |
| Chest Pain | No | 537 | 72.2 |
|  | Yes | 207 | 27.8 |
| Shortness of Breath | No | 289 | 38.8 |
|  | Yes | 455 | 61.2 |
| Cough | No | 247 | 33.4 |
|  | Yes | 492 | 66.6 |
| Diabetes Mellitus | No | 412 | 55.2 |
|  | Yes | 334 | 44.8 |
| Hypertension | No | 335 | 44.9 |
|  | Yes | 411 | 55.1 |
| Coronary Artery Disease | No | 626 | 83.9 |
|  | Yes | 120 | 16.1 |
| Heart Failure | No | 686 | 92.0 |
|  | Yes | 60 | 8.0 |
| Other Cardiovascular Diseases | No | 716 | 96.0 |
|  | Yes | 30 | 4.0 |
| Asthma | No | 706 | 94.6 |
|  | Yes | 40 | 5.4 |
| Chronic Obstructive Pulmonary Diseases | No | 723 | 96.9 |
|  | Yes | 23 | 3.1 |
| Other Respiratory Diseases | No | 674 | 90.3 |
|  | Yes | 72 | 9.7 |
| Autoimmune Diseases | No | 703 | 94.2 |
|  | Yes | 43 | 5.8 |
| Chronic Kidney Diseases | No | 653 | 87.5 |
|  | Yes | 93 | 12.5 |
| Dyslipidemia | No | 729 | 97.7 |
|  | Yes | 17 | 2.3 |
| Cancer | No | 713 | 95.6 |
|  | Yes | 33 | 4.4 |
| Neurologic Diseases | No | 702 | 94.1 |
|  | Yes | 44 | 5.9 |
| Pregnancy | No | 356 | 98.1 |
|  | Yes | 7 | 1.9 |
| Vaccination Status | Not vaccinated | 721 | 96.8 |
|  | Vaccinated | 24 | 3.2 |
| Wave | First | 265 | 35.6 |
|  | Second | 480 | 64.4 |
| Admission | Floor | 524 | 71.7 |
|  | ICU | 207 | 28.3 |
| Outcome | Discharge | 547 | 74.7 |
|  | COVID-19 related death | 171 | 23.0 |
|  | Non COVID-19 related death | 14 | 2.3 |
| Outcome among ICU patients | Discharge | 97 | 46.9 |
|  | COVID-19 related death | 110 | 53.1 |
| Variable | Mean | Standard Deviation |  |
| Age | 63.149 | 15.99 |  |
| Length of Hospital Stay | 10.09 | 9.08 |  |

### Investigations and Treatment Methods

The O2 saturation on admission was 90%-94% in 24.4% of the patients and <90% in 41.9% of them while only 22.5% of the patients had documented fever. The most frequently used imaging methods were chest x-ray, High Resolution Computed Tomography (HRCT) and Computed Tomography Pulmonary Angiography (CTPA). The most frequently observed changes on chest x-ray was bilateral infiltrates (70.9%) whereas, the most frequently observed changes on HRCT was ground glass opacifications (70.0%). Additionally, 14.0% had a positive CTPA indicating Pulmonary Embolism (PE). Regarding the laboratory investigations, troponin and D-dimer were positive in 21.1% and 85.7% of the patients, respectively. Additionally, creatinine was higher than the cut-off point for the definition of Acute Kidney Injury (AKI) in 30.5% of the patients. Whereas, the mean and standard deviation of hemoglobin (Hb) and white blood cells (WBC) count were 12.45±2.21 and 10.84E3±39.31E3, respectively. Moreover, the mean and standard deviation of the ferritin and lactate dehydrogenase (LDH) were 593.25±583.143 and 701.91±633.93, respectively. The most frequently used O2 delivery device was nasal cannula as it was used in 49.7% of the patients. The second and the third most frequently used O2 delivery devices were the simple face mask (27.3%) and non-rebreather mask (24.5%), respectively. Moreover, regarding medications used in treating COVID-19 patients, systemic corticosteroids were used in 88.6% of the patients, remdesivir in 24.6% and tocilizumab in 9.1% of them. The detailed data about the investigations and treatment methods of the patients are described in Table 2.

**Table 2.**
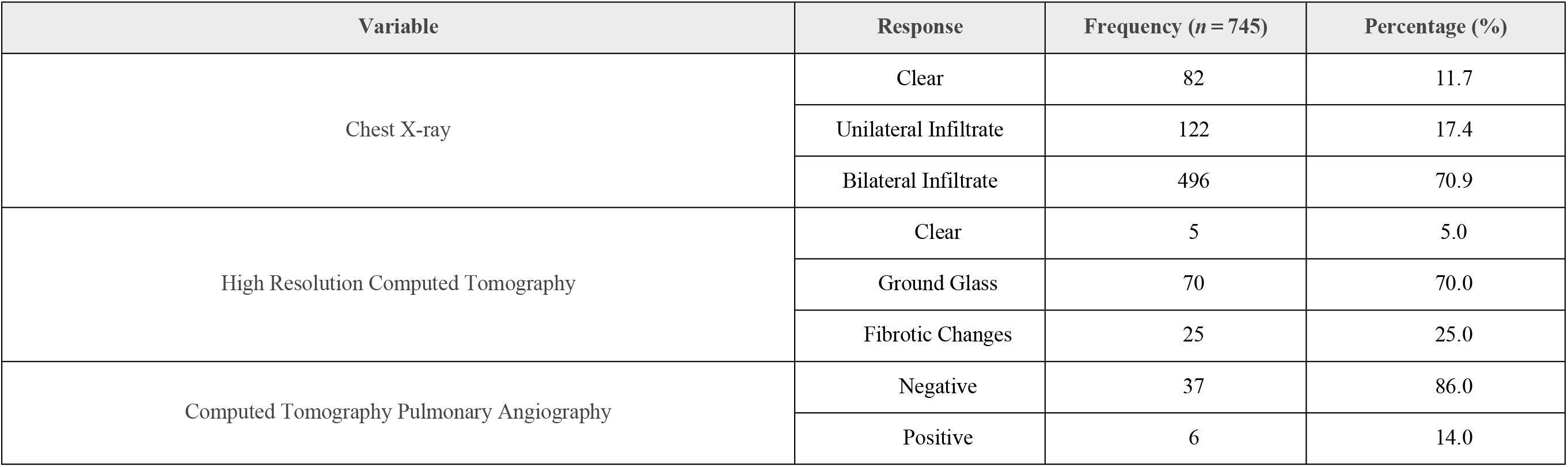
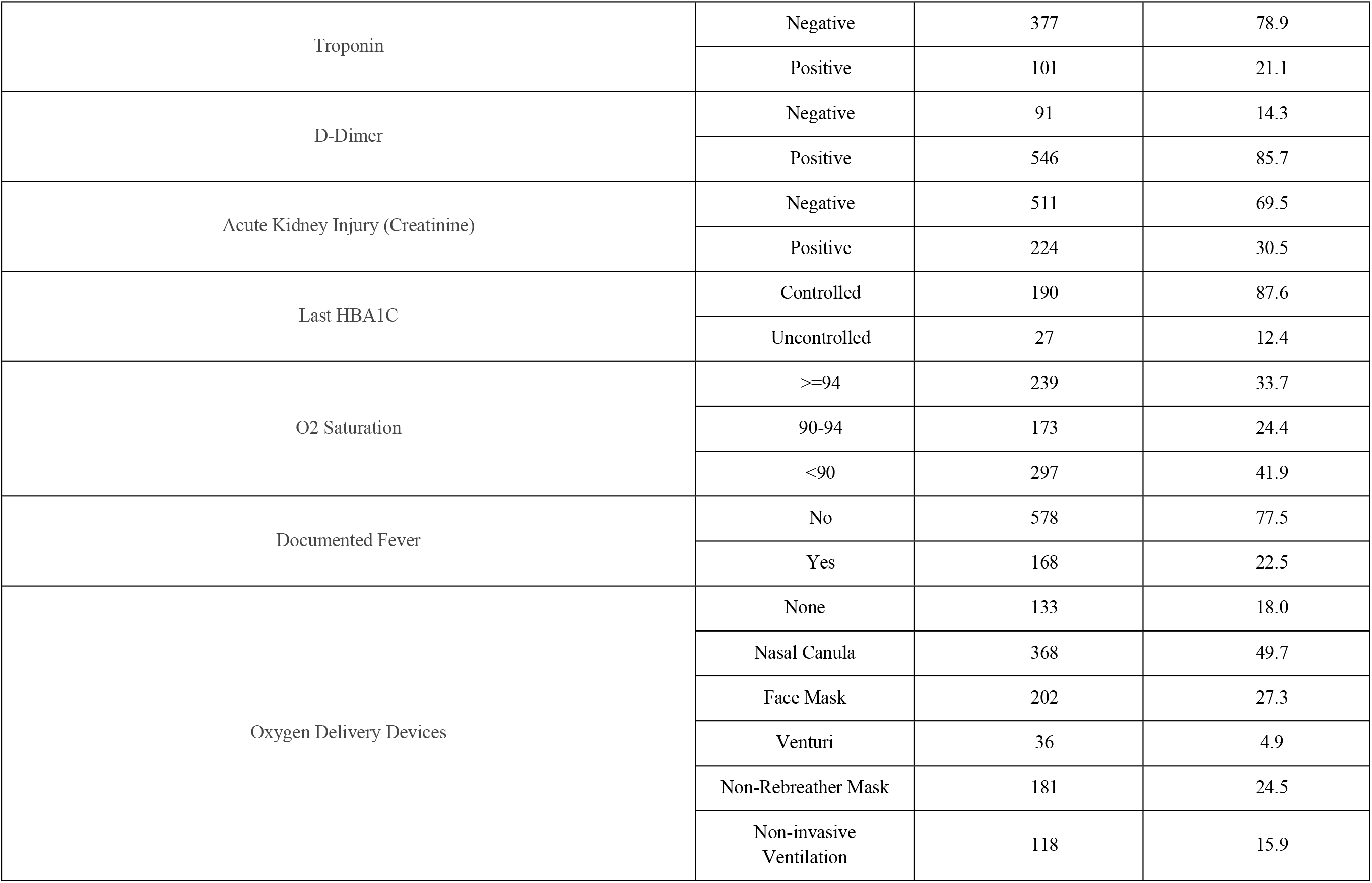
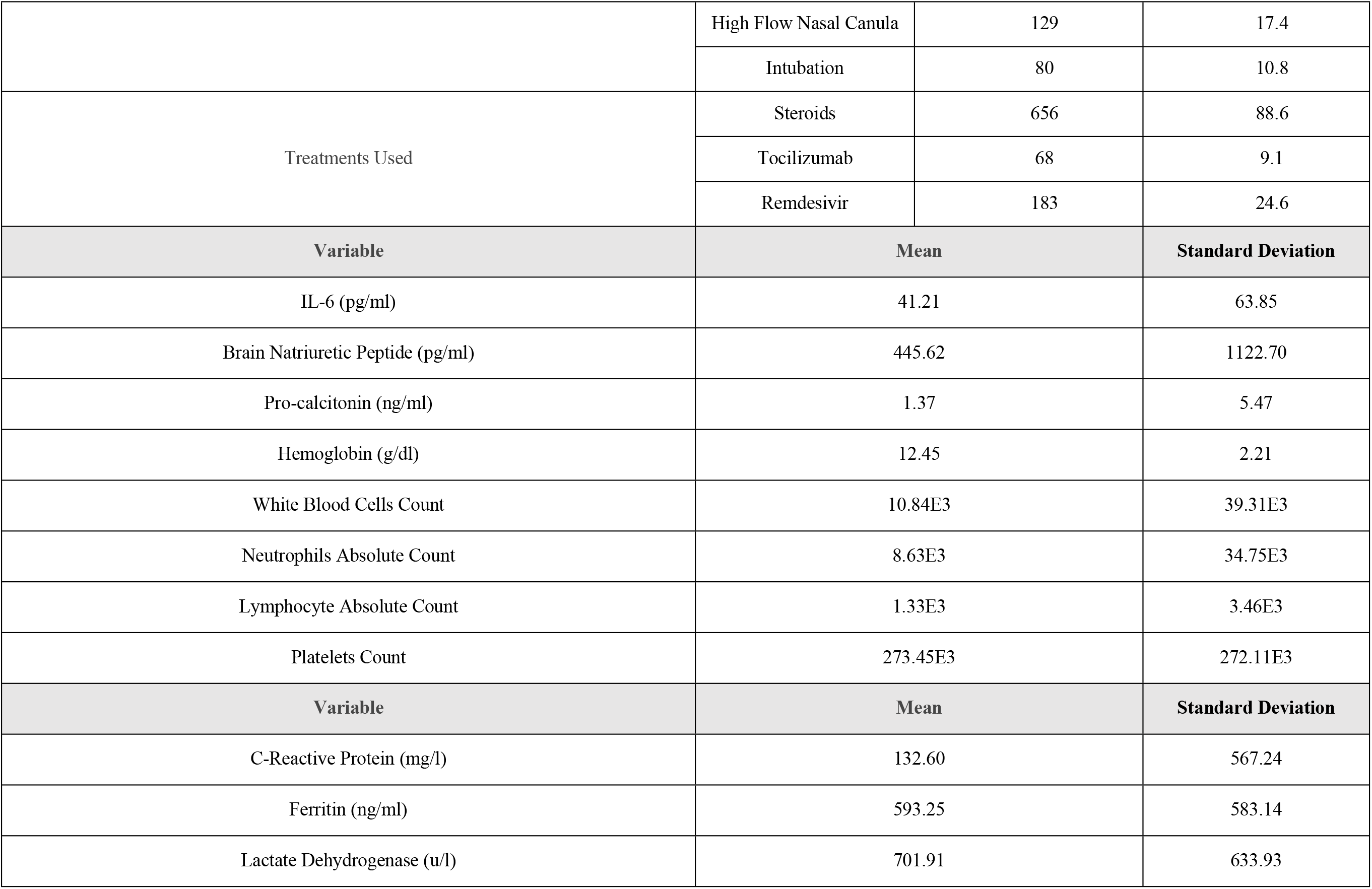
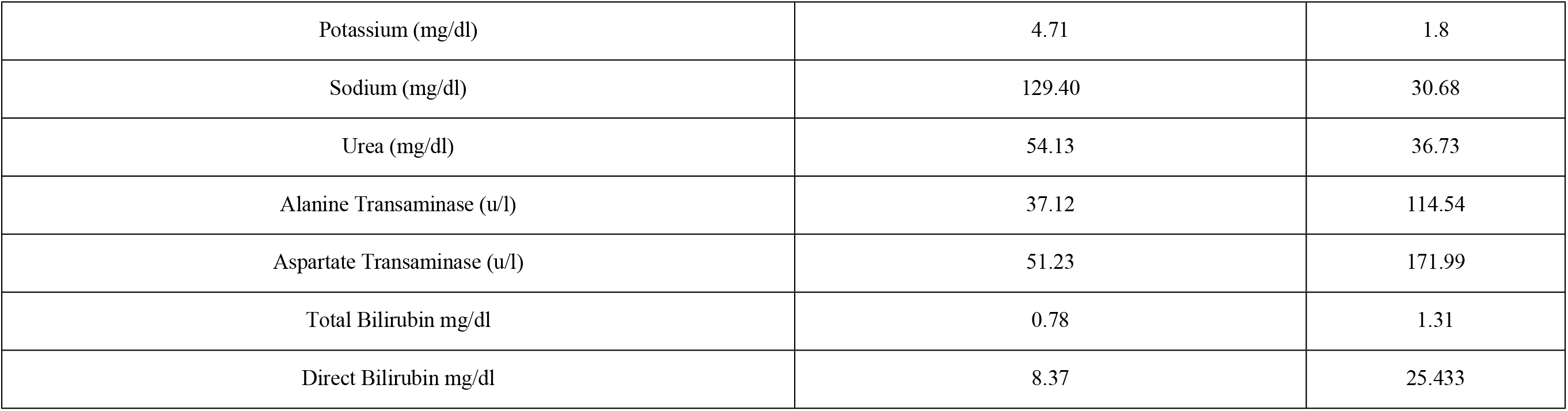
The Investigations and Treatment methods of the Patients.

| Variable | Response | Frequency (*n* = 745) | Percentage (%) |
| --- | --- | --- | --- |
| Chest X-ray | Clear | 82 | 11.7 |
|  | Unilateral Infiltrate | 122 | 17.4 |
|  | Bilateral Infiltrate | 496 | 70.9 |
| High Resolution Computed Tomography | Clear | 5 | 5.0 |
|  | Ground Glass | 70 | 70.0 |
|  | Fibrotic Changes | 25 | 25.0 |
| Computed Tomography Pulmonary Angiography | Negative | 37 | 86.0 |
|  | Positive | 6 | 14.0 |
| Troponin | Negative | 377 | 78.9 |
|  | Positive | 101 | 21.1 |
| D-Dimer | Negative | 91 | 14.3 |
|  | Positive | 546 | 85.7 |
| Acute Kidney Injury (Creatinine) | Negative | 511 | 69.5 |
|  | Positive | 224 | 30.5 |
| Last HBA1C | Controlled | 190 | 87.6 |
|  | Uncontrolled | 27 | 12.4 |
| O2 Saturation | >=94 | 239 | 33.7 |
|  | 90-94 | 173 | 24.4 |
|  | <90 | 297 | 41.9 |
| Documented Fever | No | 578 | 77.5 |
|  | Yes | 168 | 22.5 |
| Oxygen Delivery Devices | None | 133 | 18.0 |
|  | Nasal Canula | 368 | 49.7 |
|  | Face Mask | 202 | 27.3 |
|  | Venturi | 36 | 4.9 |
|  | Non-Rebreather Mask | 181 | 24.5 |
|  | Non-invasive Ventilation | 118 | 15.9 |

|  | High Flow Nasal Canula | 129 | 17.4 |
| --- | --- | --- | --- |
|  | Intubation | 80 | 10.8 |
| Treatments Used | Steroids | 656 | 88.6 |
|  | Tocilizumab | 68 | 9.1 |
|  | Remdesivir | 183 | 24.6 |
| Variable | Mean |  | Standard Deviation |
| IL-6 (pg/ml) | 41.21 |  | 63.85 |
| Brain Natriuretic Peptide (pg/ml) | 445.62 |  | 1122.70 |
| Pro-calcitonin (ng/ml) | 1.37 |  | 5.47 |
| Hemoglobin (g/dl) | 12.45 |  | 2.21 |
| White Blood Cells Count | 10.84E3 |  | 39.31E3 |
| Neutrophils Absolute Count | 8.63E3 |  | 34.75E3 |
| Lymphocyte Absolute Count | 1.33E3 |  | 3.46E3 |
| Platelets Count | 273.45E3 |  | 272.11E3 |
| Variable | Mean |  | Standard Deviation |
| C-Reactive Protein (mg/l) | 132.60 |  | 567.24 |
| Ferritin (ng/ml) | 593.25 |  | 583.14 |
| Lactate Dehydrogenase (u/l) | 701.91 |  | 633.93 |

|  |  |  |
| --- | --- | --- |
| Potassium (mg/dl) | 4.71 | 1.8 |
| Sodium (mg/dl) | 129.40 | 30.68 |
| Urea (mg/dl) | 54.13 | 36.73 |
| Alanine Transaminase (u/l) | 37.12 | 114.54 |
| Aspartate Transaminase (u/l) | 51.23 | 171.99 |
| Total Bilirubin mg/dl | 0.78 | 1.31 |
| Direct Bilirubin mg/dl | 8.37 | 25.433 |

### Predictors of COVID-19 Outcomes

Regarding the outcomes of COVID-19 patients, 23.0% of all the patients died (COVID-19 related death) while 53.1% of the patients who were admitted to ICU died (Table 1). In the non-adjusted model, age, DM, HTN, CAD,HF CKD, neurologic diseases, chest x-ray patterns, troponin, hemoglobin, WBCs, neutrophils, D-dimer, AKI, LDH, Potassium, total bilirubin and O2 saturation <90% were significant predictors for mortality. After adjustment for confounding variables, CKD (AOR=3.831;95% CI: 1.179-12.446), troponin (AOR=3.060;95% CI: 1.156-8.102), LDH (AOR=1.002;95% CI: 1.001-1.003) and O2 saturation <90% (AOR=2.761;95% CI: 1.066-7.155) were the only significant predictors for mortality (Table 3). Patients who had CKD, positive troponin, high LDH values and O2 saturation<90% had a significantly higher risk of mortality. Additionally, regarding COVID-19 severity, 28.3% of the patients were admitted to the ICU and 10.8% of the patients were intubated (Table 1). In the non-adjusted model that investigated ICU admission, age, smoking status, GI symptoms, DM, HTN, CKD, HF, other CVDs, chest x-ray patterns, troponin, GI symptoms, neutrophils, D-dimer, AKI, potassium, sodium, O2 saturation <90%, Tocilizumab use and remdesivir use were significant predictors for ICU admission. However, in the adjusted model, only HF (AOR=7.894;95% CI: 1.391-44.818) and remdesivir use (AOR=0.192;95% CI: 0.044-0.835) were significant predictors for ICU admission (Table 3). Patients who had HF had significantly higher risk for ICU admission while patients who were treated using remdesivir had significantly lower risk for ICU admission. Furthermore, in the non-adjusted model that evaluated invasive intubation, GI symptoms, CKD, neurological diseases, chest x-ray patterns, troponin, D-dimer, AKI, sodium, ALT, AST, O2 saturation <90% and steroids were significant predictors for invasive intubation. In the adjusted model, O2 saturation<90% (AOR=16.585;95% CI: 2.892-95.118) and GI symptoms (AOR=0.323;95% CI: 0.105-0.993) were the only significant predictors for invasive intubation (Table 3). Patients who had O2 saturation <90% had significantly higher risk for invasive intubation whereas patients who reported GI symptoms had significantly lower risk for it. In regards to the length of hospital stay, the mean and standard deviation for hospital stay was 10.09 ± 9.08 (Table 1). In the non-adjusted model, pregnancy, chest x-ray patterns, Hb, D-dimer, total bilirubin, O2 saturation, Tocilizumab use and remdesivir use were significant predictors for the length of hospital stay. However, after adjusting for confounding variables, none of the aforementioned variables were significant predictors for the length of hospital stay (Table 3).

**Table 3.**
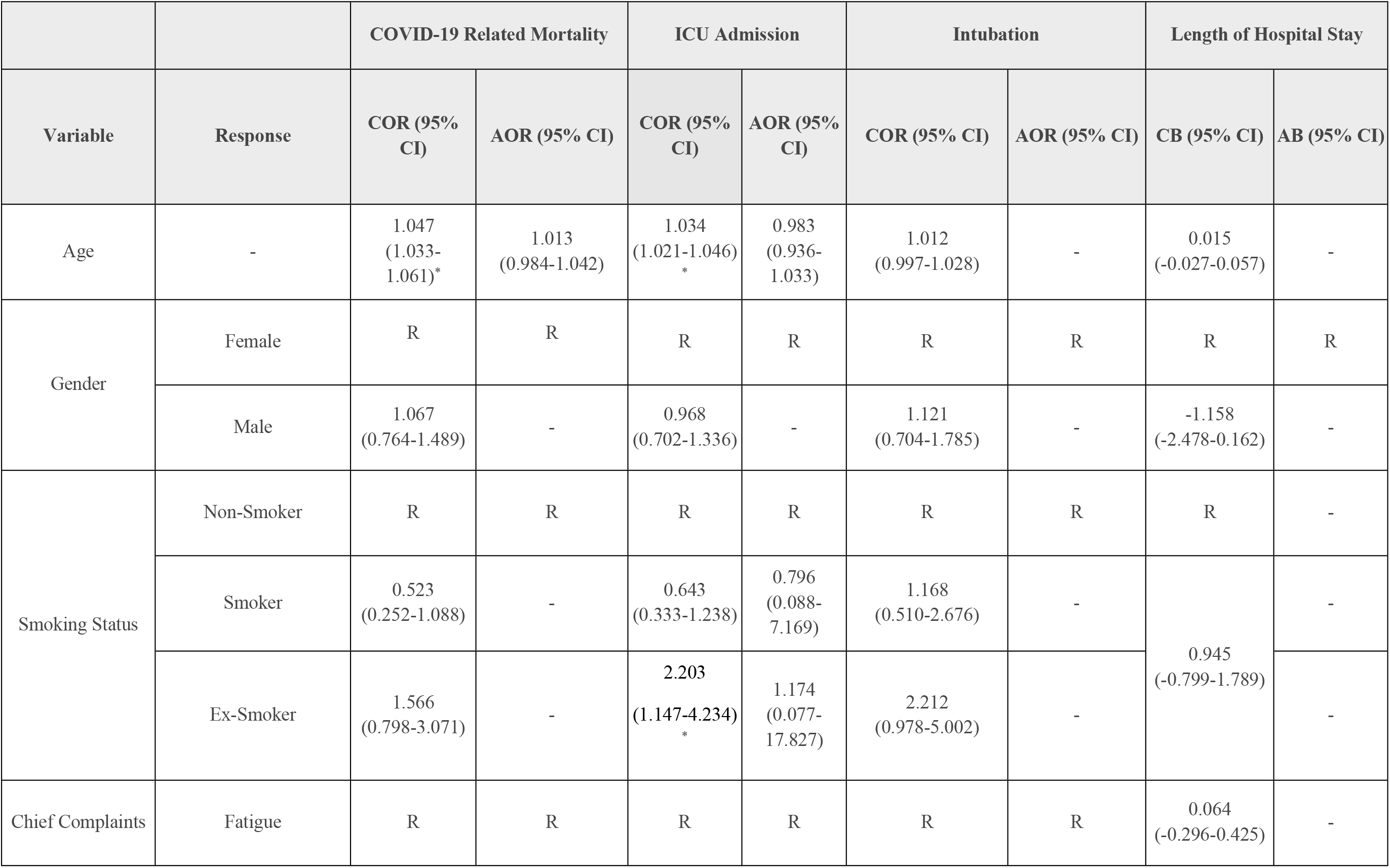
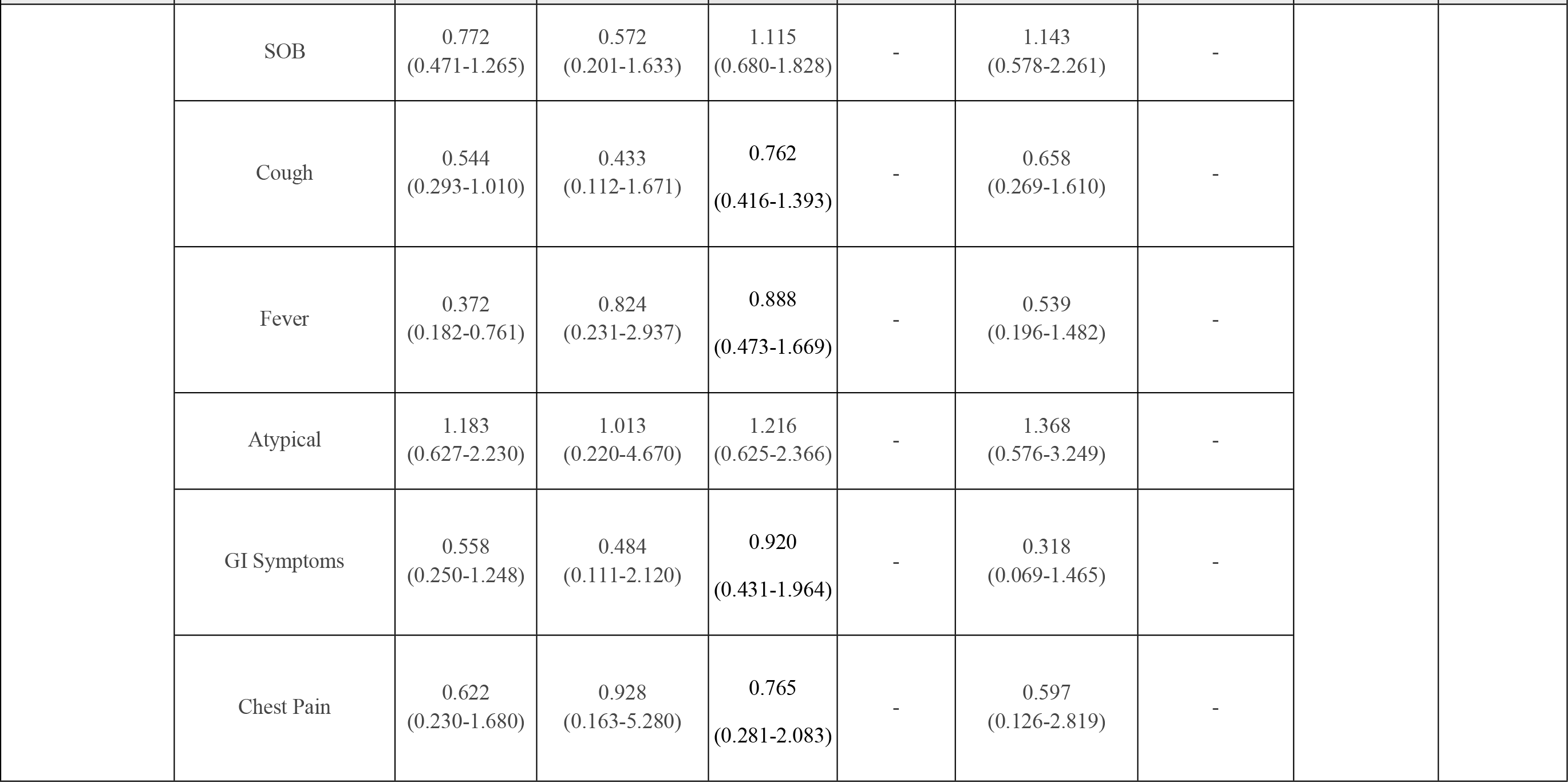
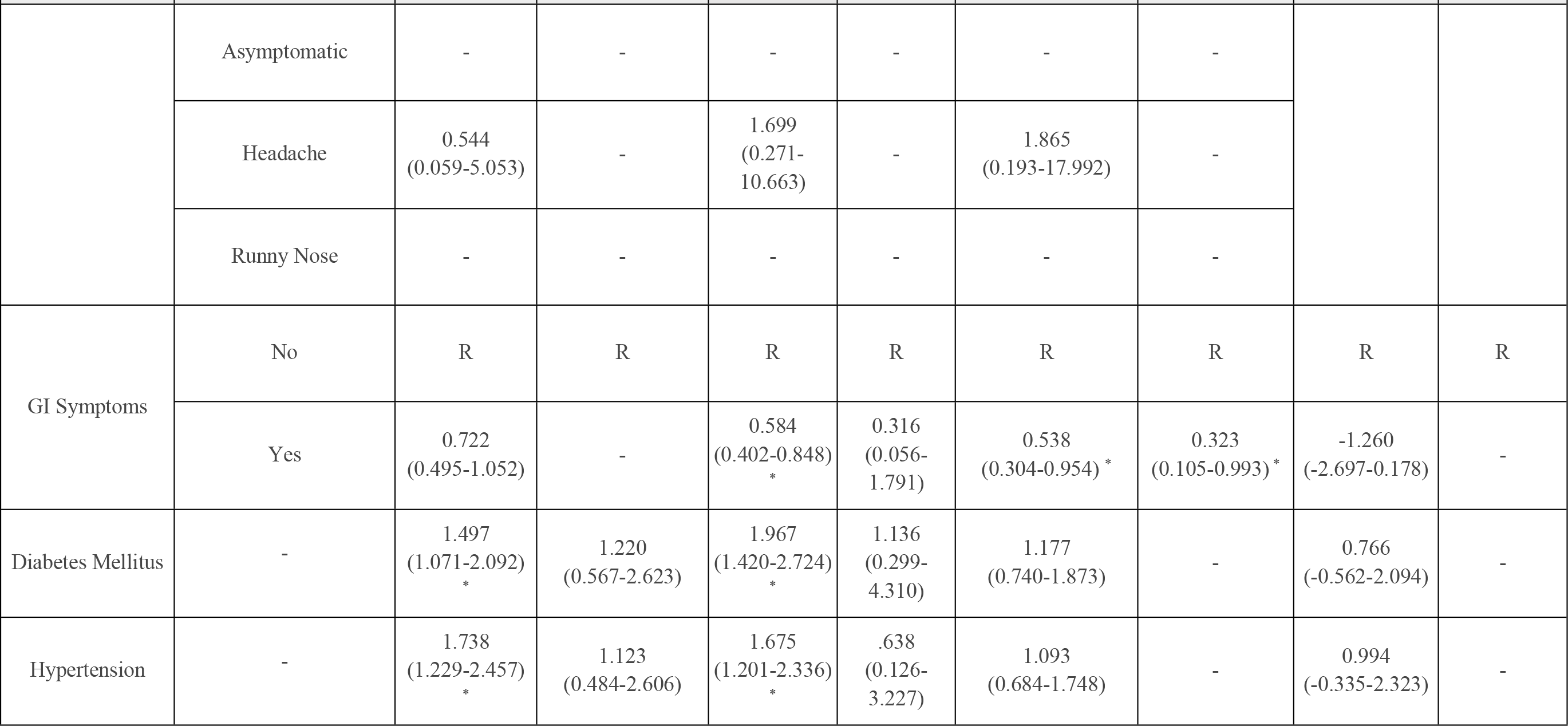
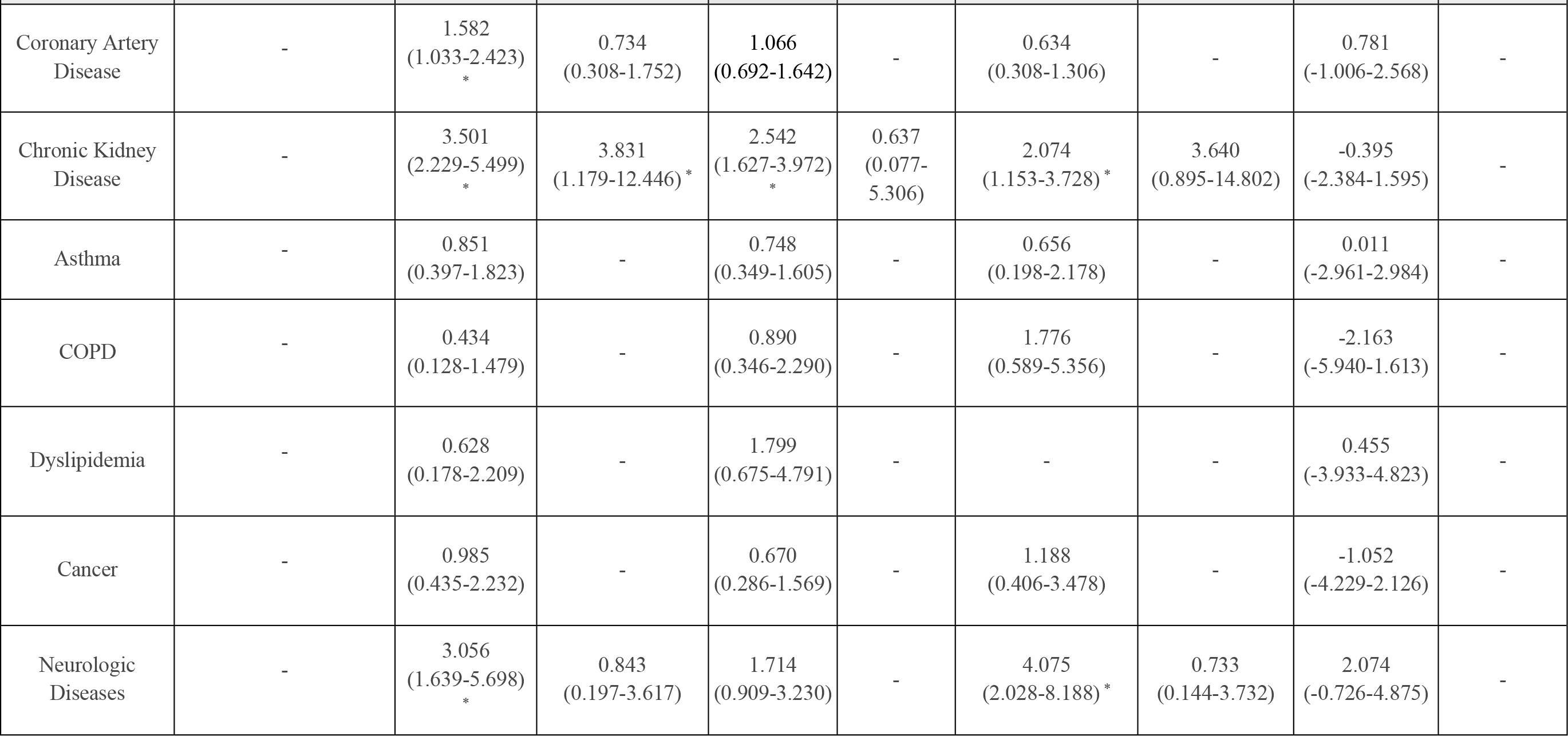
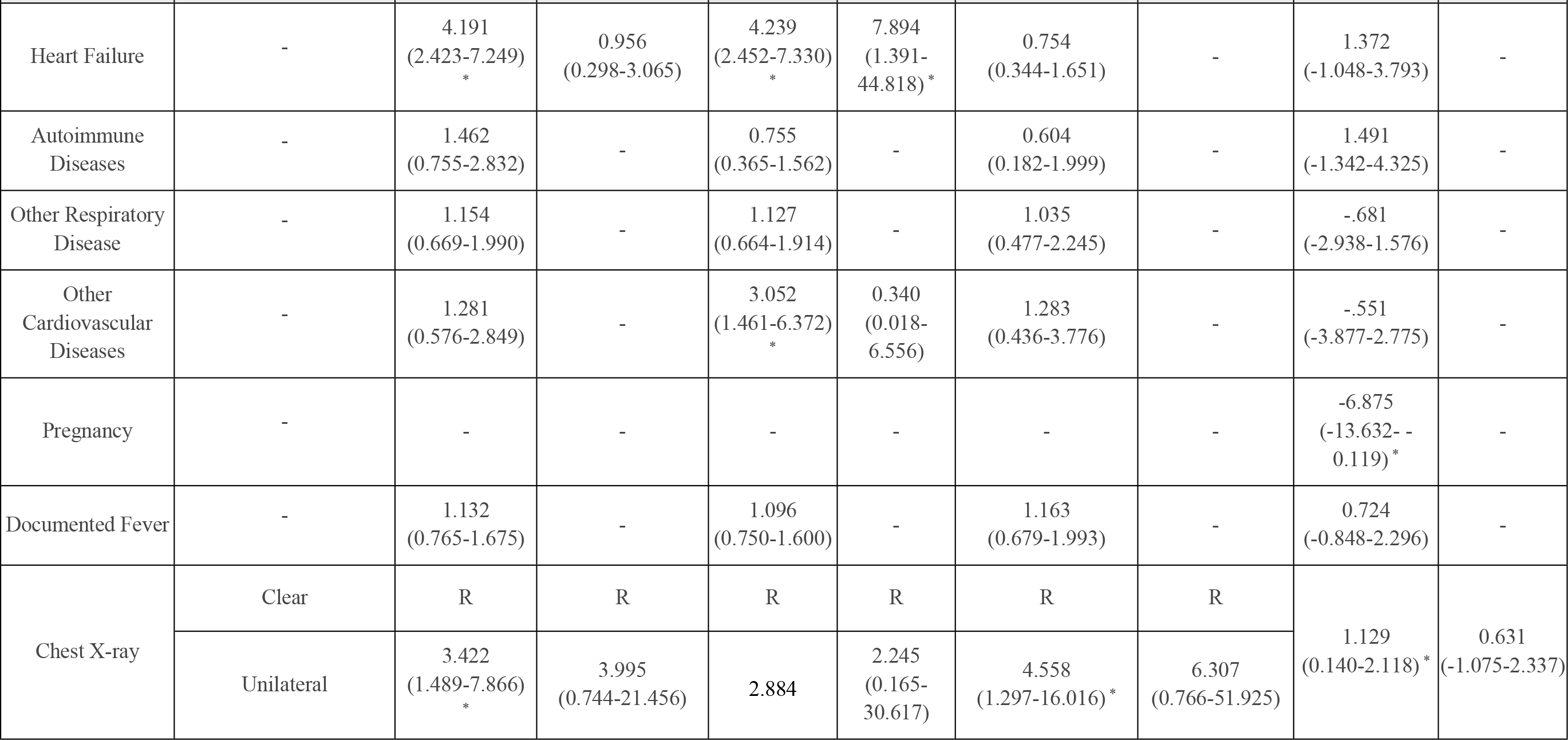
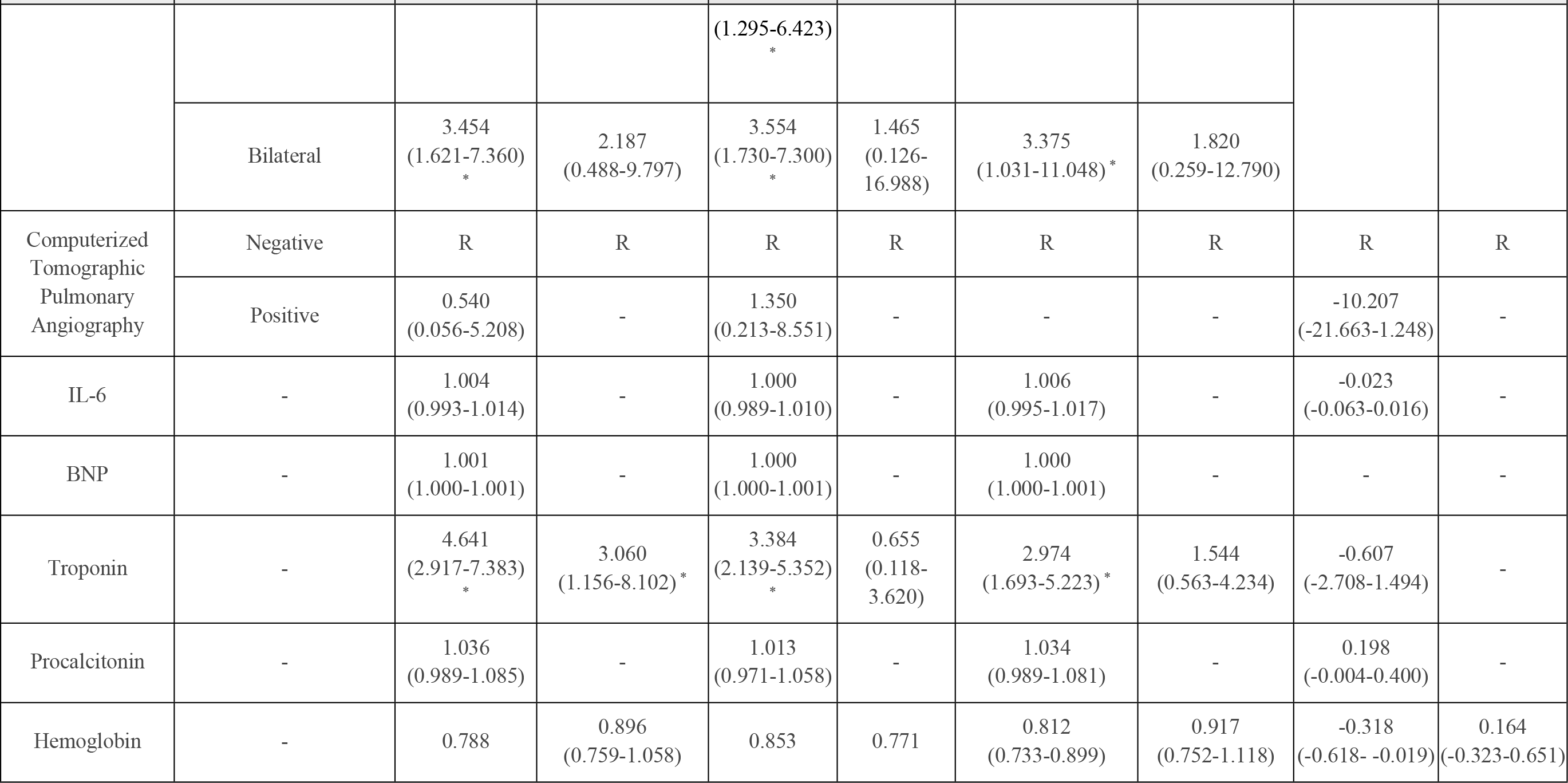
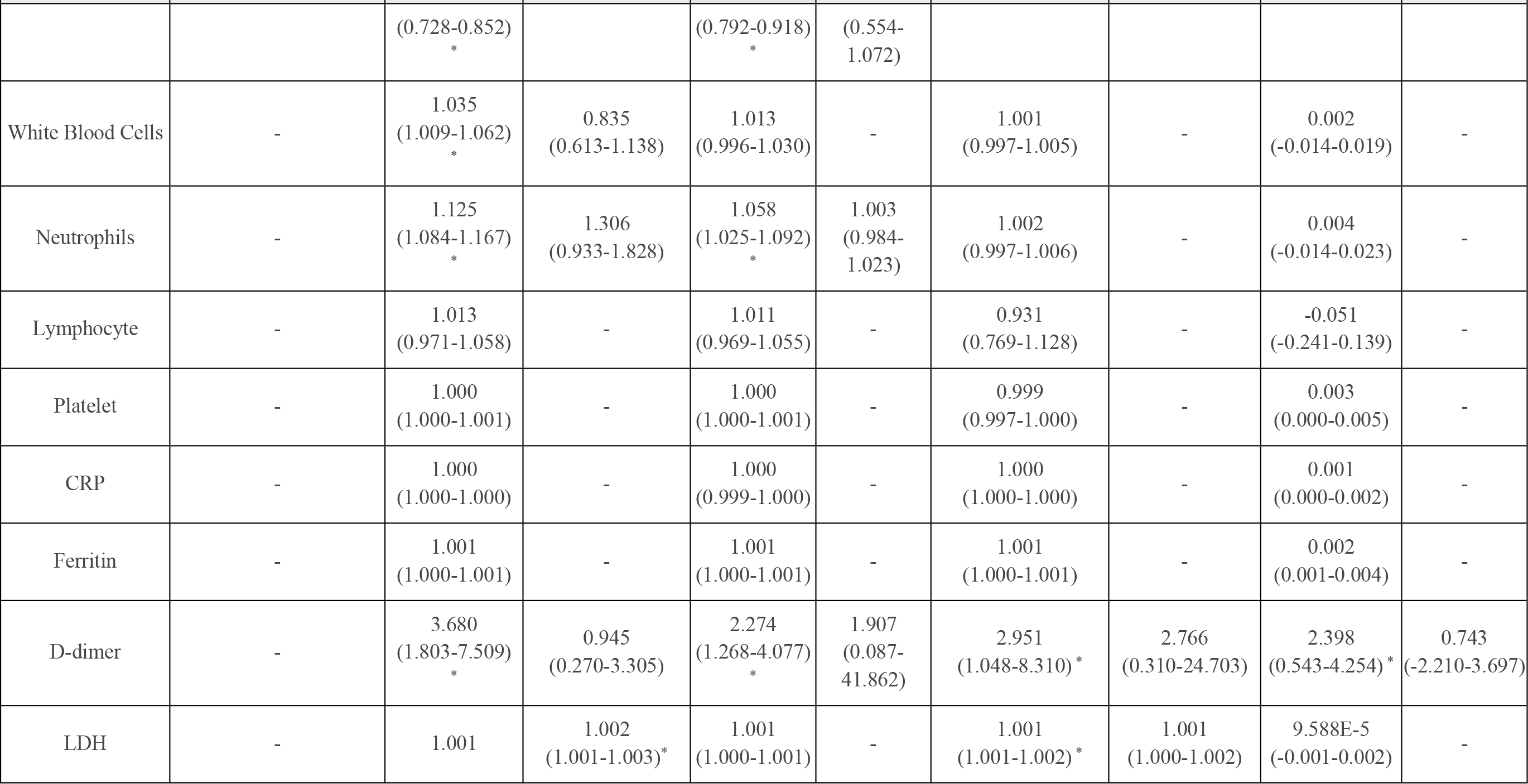
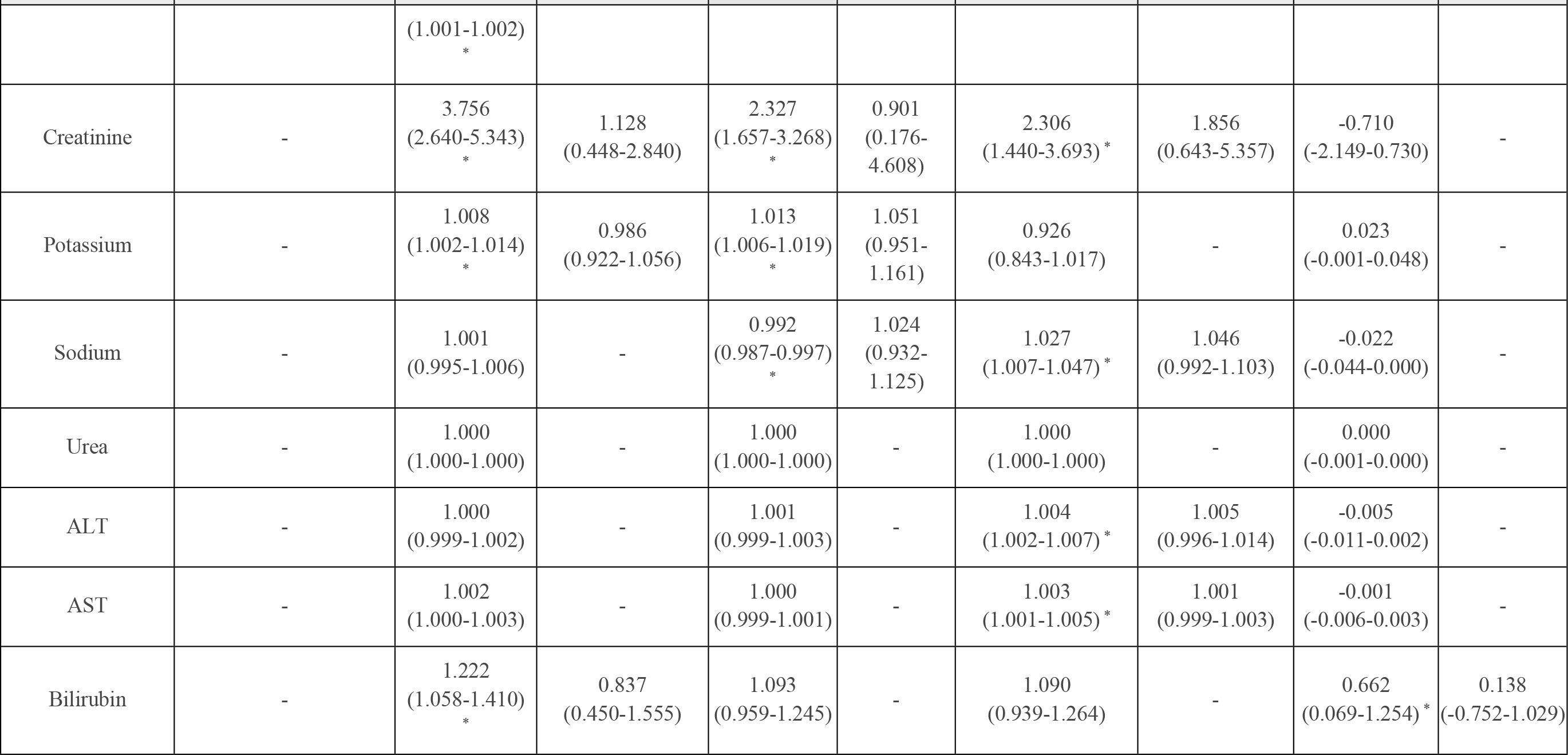
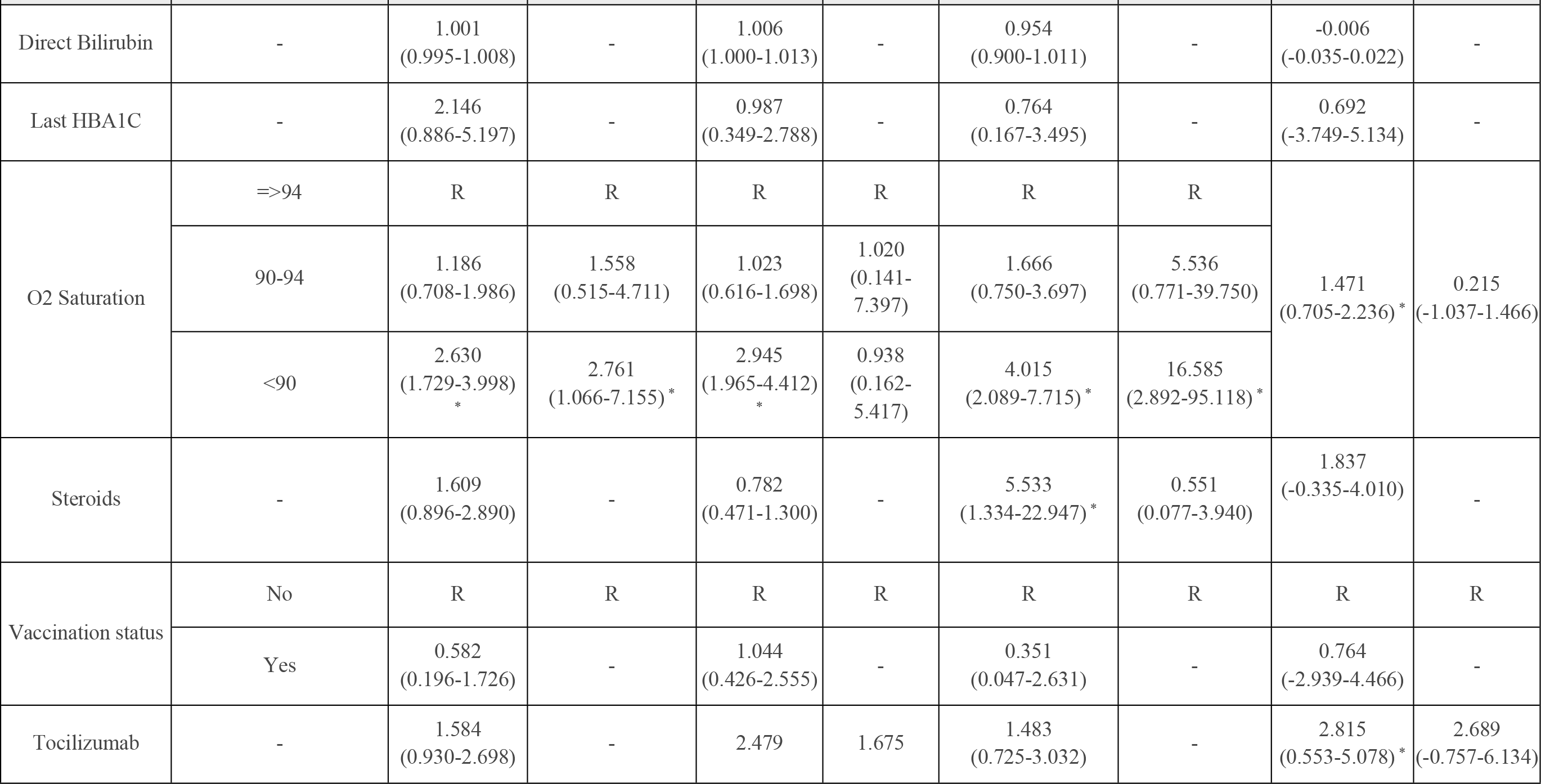
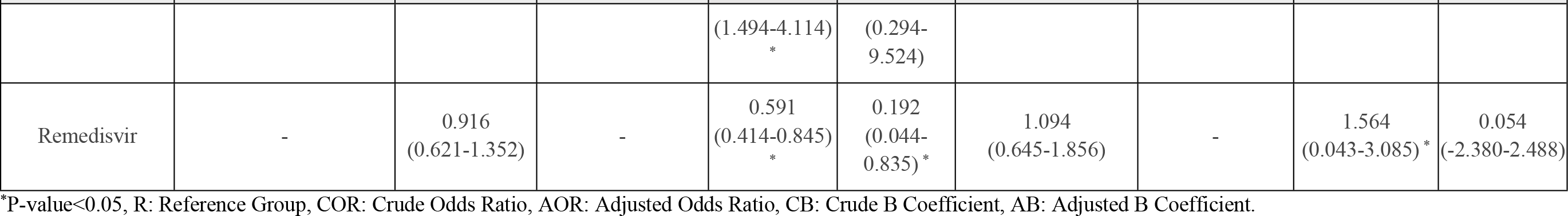
Regression Analysis for the Predictors of COVID-19 Outcomes.

|  |  | COVID-19 Related Mortality |  | ICU Admission |  | Intubation |  | Length of Hospital Stay |  |
| --- | --- | --- | --- | --- | --- | --- | --- | --- | --- |
| Variable | Response | COR (95% CI) | AOR (95% CI) | COR (95% CI) | AOR (95% CI) | COR (95% CI) | AOR (95% CI) | CB (95% CI) | AB (95% CI) |
| Age | - | 1.047<br>(1.033-1.061)* | 1.013<br>(0.984-1.042) | 1.034<br>(1.021-1.046)<br>* | 0.983<br>(0.936-1.033) | 1.012<br>(0.997-1.028) | - | 0.015<br>(-0.027-0.057) | - |
| Gender | Female | R | R | R | R | R | R | R | R |
|  | Male | 1.067<br>(0.764-1.489) | - | 0.968<br>(0.702-1.336) | - | 1.121<br>(0.704-1.785) | - | -1.158<br>(-2.478-0.162) | - |
| Smoking Status | Non-Smoker | R | R | R | R | R | R | R | - |
|  | Smoker | 0.523<br>(0.252-1.088) | - | 0.643<br>(0.333-1.238) | 0.796<br>(0.088-7.169) | 1.168<br>(0.510-2.676) | - | 0.945<br>(-0.799-1.789) | - |
|  | Ex-Smoker | 1.566<br>(0.798-3.071) | - | 2.203<br>(1.147-4.234)<br>* | 1.174<br>(0.077-17.827) | 2.212<br>(0.978-5.002) | - |  | - |
| Chief Complaints | Fatigue | R | R | R | R | R | R | 0.064<br>(-0.296-0.425) | - |
| Variable | Response | COR (95% CI) | AOR (95% CI) | COR (95% CI) | AOR (95% CI) | COR (95% CI) | AOR (95% CI) | CB (95% CI) | AB (95% CI) |
|  | SOB | 0.772<br>(0.471-1.265) | 0.572<br>(0.201-1.633) | 1.115<br>(0.680-1.828) | - | 1.143<br>(0.578-2.261) | - |  |  |
|  | Cough | 0.544<br>(0.293-1.010) | 0.433<br>(0.112-1.671) | 0.762<br>(0.416-1.393) | - | 0.658<br>(0.269-1.610) | - |  |  |
|  | Fever | 0.372<br>(0.182-0.761) | 0.824<br>(0.231-2.937) | 0.888<br>(0.473-1.669) | - | 0.539<br>(0.196-1.482) | - |  |  |
|  | Atypical | 1.183<br>(0.627-2.230) | 1.013<br>(0.220-4.670) | 1.216<br>(0.625-2.366) | - | 1.368<br>(0.576-3.249) | - |  |  |
|  | GI Symptoms | 0.558<br>(0.250-1.248) | 0.484<br>(0.111-2.120) | 0.920<br>(0.431-1.964) | - | 0.318<br>(0.069-1.465) | - |  |  |
|  | Chest Pain | 0.622<br>(0.230-1.680) | 0.928<br>(0.163-5.280) | 0.765<br>(0.281-2.083) | - | 0.597<br>(0.126-2.819) | - |  |  |
| Variable | Response | COR (95% CI) | AOR (95% CI) | COR (95% CI) | AOR (95% CI) | COR (95% CI) | AOR (95% CI) | CB (95% CI) | AB (95% CI) |
|  | Asymptomatic | - | - | - | - | - | - |  |  |
|  | Headache | 0.544<br>(0.059-5.053) | - | 1.699<br>(0.271-10.663) | - | 1.865<br>(0.193-17.992) | - |  |  |
|  | Runny Nose | - | - | - | - | - | - |  |  |
| GI Symptoms | No | R | R | R | R | R | R | R | R |
|  | Yes | 0.722<br>(0.495-1.052) | - | 0.584<br>(0.402-0.848)<br>* | 0.316<br>(0.056-1.791) | 0.538<br>(0.304-0.954) * | 0.323<br>(0.105-0.993) * | -1.260<br>(-2.697-0.178) | - |
| Diabetes Mellitus | - | 1.497<br>(1.071-2.092)<br>* | 1.220<br>(0.567-2.623) | 1.967<br>(1.420-2.724)<br>* | 1.136<br>(0.299-4.310) | 1.177<br>(0.740-1.873) | - | 0.766<br>(-0.562-2.094) | - |
| Hypertension | - | 1.738<br>(1.229-2.457)<br>* | 1.123<br>(0.484-2.606) | 1.675<br>(1.201-2.336)<br>* | .638<br>(0.126-3.227) | 1.093<br>(0.684-1.748) | - | 0.994<br>(-0.335-2.323) | - |
| Variable | Response | COR (95% CI) | AOR (95% CI) | COR (95% CI) | AOR (95% CI) | COR (95% CI) | AOR (95% CI) | CB (95% CI) | AB (95% CI) |
| Coronary Artery Disease | - | 1.582<br>(1.033-2.423)<br>* | 0.734<br>(0.308-1.752) | 1.066<br>(0.692-1.642) | - | 0.634<br>(0.308-1.306) | - | 0.781<br>(-1.006-2.568) | - |
| Chronic Kidney Disease | - | 3.501<br>(2.229-5.499)<br>* | 3.831<br>(1.179-12.446) * | 2.542<br>(1.627-3.972)<br>* | 0.637<br>(0.077-5.306) | 2.074<br>(1.153-3.728) * | 3.640<br>(0.895-14.802) | -0.395<br>(-2.384-1.595) | - |
| Asthma | - | 0.851<br>(0.397-1.823) | - | 0.748<br>(0.349-1.605) | - | 0.656<br>(0.198-2.178) | - | 0.011<br>(-2.961-2.984) | - |
| COPD | - | 0.434<br>(0.128-1.479) | - | 0.890<br>(0.346-2.290) | - | 1.776<br>(0.589-5.356) | - | -2.163<br>(-5.940-1.613) | - |
| Dyslipidemia | - | 0.628<br>(0.178-2.209) | - | 1.799<br>(0.675-4.791) | - | - | - | 0.455<br>(-3.933-4.823) | - |
| Cancer | - | 0.985<br>(0.435-2.232) | - | 0.670<br>(0.286-1.569) | - | 1.188<br>(0.406-3.478) | - | -1.052<br>(-4.229-2.126) | - |
| Neurologic Diseases | - | 3.056<br>(1.639-5.698)<br>* | 0.843<br>(0.197-3.617) | 1.714<br>(0.909-3.230) | - | 4.075<br>(2.028-8.188) * | 0.733<br>(0.144-3.732) | 2.074<br>(-0.726-4.875) | - |
| Variable | Response | COR (95% CI) | AOR (95% CI) | COR (95% CI) | AOR (95% CI) | COR (95% CI) | AOR (95% CI) | CB (95% CI) | AB (95% CI) |
| Heart Failure | - | 4.191<br>(2.423-7.249)<br>* | 0.956<br>(0.298-3.065) | 4.239<br>(2.452-7.330)<br>* | 7.894<br>(1.391-44.818)* | 0.754<br>(0.344-1.651) | - | 1.372<br>(-1.048-3.793) | - |
| Autoimmune Diseases | - | 1.462<br>(0.755-2.832) | - | 0.755<br>(0.365-1.562) | - | 0.604<br>(0.182-1.999) | - | 1.491<br>(-1.342-4.325) | - |
| Other Respiratory Disease | - | 1.154<br>(0.669-1.990) | - | 1.127<br>(0.664-1.914) | - | 1.035<br>(0.477-2.245) | - | -.681<br>(-2.938-1.576) | - |
| Other Cardiovascular Diseases | - | 1.281<br>(0.576-2.849) | - | 3.052<br>(1.461-6.372)<br>* | 0.340<br>(0.018-6.556) | 1.283<br>(0.436-3.776) | - | -.551<br>(-3.877-2.775) | - |
| Pregnancy | - | - | - | - | - | - | - | -6.875<br>(-13.632- -0.119)* | - |
| Documented Fever | - | 1.132<br>(0.765-1.675) | - | 1.096<br>(0.750-1.600) | - | 1.163<br>(0.679-1.993) | - | 0.724<br>(-0.848-2.296) | - |
| Chest X-ray | Clear | R | R | R | R | R | R | 1.129<br>(0.140-2.118)* | 0.631<br>(-1.075-2.337) |
|  | Unilateral | 3.422<br>(1.489-7.866)<br>* | 3.995<br>(0.744-21.456) | 2.884 | 2.245<br>(0.165-30.617) | 4.558<br>(1.297-16.016)* | 6.307<br>(0.766-51.925) |  |  |
| Variable | Response | COR (95% CI) | AOR (95% CI) | COR (95% CI) | AOR (95% CI) | COR (95% CI) | AOR (95% CI) | CB (95% CI) | AB (95% CI) |
|  |  |  |  | (1.295-6.423)<br>* |  |  |  |  |  |
|  | Bilateral | 3.454<br>(1.621-7.360)<br>* | 2.187<br>(0.488-9.797) | 3.554<br>(1.730-7.300)<br>* | 1.465<br>(0.126-16.988) | 3.375<br>(1.031-11.048) * | 1.820<br>(0.259-12.790) |  |  |
| Computerized Tomographic Pulmonary Angiography | Negative | R | R | R | R | R | R | R | R |
|  | Positive | 0.540<br>(0.056-5.208) | - | 1.350<br>(0.213-8.551) | - | - | - | -10.207<br>(-21.663-1.248) | - |
| IL-6 | - | 1.004<br>(0.993-1.014) | - | 1.000<br>(0.989-1.010) | - | 1.006<br>(0.995-1.017) | - | -0.023<br>(-0.063-0.016) | - |
| BNP | - | 1.001<br>(1.000-1.001) | - | 1.000<br>(1.000-1.001) | - | 1.000<br>(1.000-1.001) | - | - | - |
| Troponin | - | 4.641<br>(2.917-7.383)<br>* | 3.060<br>(1.156-8.102) * | 3.384<br>(2.139-5.352)<br>* | 0.655<br>(0.118-3.620) | 2.974<br>(1.693-5.223) * | 1.544<br>(0.563-4.234) | -0.607<br>(-2.708-1.494) | - |
| Procalcitonin | - | 1.036<br>(0.989-1.085) | - | 1.013<br>(0.971-1.058) | - | 1.034<br>(0.989-1.081) | - | 0.198<br>(-0.004-0.400) | - |
| Hemoglobin | - | 0.788 | 0.896<br>(0.759-1.058) | 0.853 | 0.771 | 0.812<br>(0.733-0.899) | 0.917<br>(0.752-1.118) | -0.318<br>(-0.618- -0.019) | 0.164<br>(-0.323-0.651) |
| Variable | Response | COR (95% CI) | AOR (95% CI) | COR (95% CI) | AOR (95% CI) | COR (95% CI) | AOR (95% CI) | CB (95% CI) | AB (95% CI) |
|  |  | (0.728-0.852)<br>* |  | (0.792-0.918)<br>* | (0.554-1.072) |  |  |  |  |
| White Blood Cells | - | 1.035<br>(1.009-1.062)<br>* | 0.835<br>(0.613-1.138) | 1.013<br>(0.996-1.030) | - | 1.001<br>(0.997-1.005) | - | 0.002<br>(-0.014-0.019) | - |
| Neutrophils | - | 1.125<br>(1.084-1.167)<br>* | 1.306<br>(0.933-1.828) | 1.058<br>(1.025-1.092)<br>* | 1.003<br>(0.984-1.023) | 1.002<br>(0.997-1.006) | - | 0.004<br>(-0.014-0.023) | - |
| Lymphocyte | - | 1.013<br>(0.971-1.058) | - | 1.011<br>(0.969-1.055) | - | 0.931<br>(0.769-1.128) | - | -0.051<br>(-0.241-0.139) | - |
| Platelet | - | 1.000<br>(1.000-1.001) | - | 1.000<br>(1.000-1.001) | - | 0.999<br>(0.997-1.000) | - | 0.003<br>(0.000-0.005) | - |
| CRP | - | 1.000<br>(1.000-1.000) | - | 1.000<br>(0.999-1.000) | - | 1.000<br>(1.000-1.000) | - | 0.001<br>(0.000-0.002) | - |
| Ferritin | - | 1.001<br>(1.000-1.001) | - | 1.001<br>(1.000-1.001) | - | 1.001<br>(1.000-1.001) | - | 0.002<br>(0.001-0.004) | - |
| D-dimer | - | 3.680<br>(1.803-7.509)<br>* | 0.945<br>(0.270-3.305) | 2.274<br>(1.268-4.077)<br>* | 1.907<br>(0.087-41.862) | 2.951<br>(1.048-8.310)* | 2.766<br>(0.310-24.703) | 2.398<br>(0.543-4.254)* | 0.743<br>(-2.210-3.697) |
| LDH | - | 1.001 | 1.002<br>(1.001-1.003)* | 1.001<br>(1.000-1.001) | - | 1.001<br>(1.001-1.002)* | 1.001<br>(1.000-1.002) | 9.588E-5<br>(-0.001-0.002) | - |
| Variable | Response | COR (95% CI) | AOR (95% CI) | COR (95% CI) | AOR (95% CI) | COR (95% CI) | AOR (95% CI) | CB (95% CI) | AB (95% CI) |
|  |  | (1.001-1.002)<br>* |  |  |  |  |  |  |  |
| Creatinine | - | 3.756<br>(2.640-5.343)<br>* | 1.128<br>(0.448-2.840) | 2.327<br>(1.657-3.268)<br>* | 0.901<br>(0.176-4.608) | 2.306<br>(1.440-3.693) * | 1.856<br>(0.643-5.357) | -0.710<br>(-2.149-0.730) | - |
| Potassium | - | 1.008<br>(1.002-1.014)<br>* | 0.986<br>(0.922-1.056) | 1.013<br>(1.006-1.019)<br>* | 1.051<br>(0.951-1.161) | 0.926<br>(0.843-1.017) | - | 0.023<br>(-0.001-0.048) | - |
| Sodium | - | 1.001<br>(0.995-1.006) | - | 0.992<br>(0.987-0.997)<br>* | 1.024<br>(0.932-1.125) | 1.027<br>(1.007-1.047) * | 1.046<br>(0.992-1.103) | -0.022<br>(-0.044-0.000) | - |
| Urea | - | 1.000<br>(1.000-1.000) | - | 1.000<br>(1.000-1.000) | - | 1.000<br>(1.000-1.000) | - | 0.000<br>(-0.001-0.000) | - |
| ALT | - | 1.000<br>(0.999-1.002) | - | 1.001<br>(0.999-1.003) | - | 1.004<br>(1.002-1.007) * | 1.005<br>(0.996-1.014) | -0.005<br>(-0.011-0.002) | - |
| AST | - | 1.002<br>(1.000-1.003) | - | 1.000<br>(0.999-1.001) | - | 1.003<br>(1.001-1.005) * | 1.001<br>(0.999-1.003) | -0.001<br>(-0.006-0.003) | - |
| Bilirubin | - | 1.222<br>(1.058-1.410)<br>* | 0.837<br>(0.450-1.555) | 1.093<br>(0.959-1.245) | - | 1.090<br>(0.939-1.264) | - | 0.662<br>(0.069-1.254) * | 0.138<br>(-0.752-1.029) |
| Variable | Response | COR (95% CI) | AOR (95% CI) | COR (95% CI) | AOR (95% CI) | COR (95% CI) | AOR (95% CI) | CB (95% CI) | AB (95% CI) |
| Direct Bilirubin | - | 1.001<br>(0.995-1.008) | - | 1.006<br>(1.000-1.013) | - | 0.954<br>(0.900-1.011) | - | -0.006<br>(-0.035-0.022) | - |
| Last HBA1C | - | 2.146<br>(0.886-5.197) | - | 0.987<br>(0.349-2.788) | - | 0.764<br>(0.167-3.495) | - | 0.692<br>(-3.749-5.134) | - |
| O2 Saturation | =>94 | R | R | R | R | R | R | 1.471<br>(0.705-2.236) * | 0.215<br>(-1.037-1.466) |
|  | 90-94 | 1.186<br>(0.708-1.986) | 1.558<br>(0.515-4.711) | 1.023<br>(0.616-1.698) | 1.020<br>(0.141-7.397) | 1.666<br>(0.750-3.697) | 5.536<br>(0.771-39.750) |  |  |
|  | <90 | 2.630<br>(1.729-3.998)<br>* | 2.761<br>(1.066-7.155) * | 2.945<br>(1.965-4.412)<br>* | 0.938<br>(0.162-5.417) | 4.015<br>(2.089-7.715) * | 16.585<br>(2.892-95.118) * |  |  |
| Steroids | - | 1.609<br>(0.896-2.890) | - | 0.782<br>(0.471-1.300) | - | 5.533<br>(1.334-22.947) * | 0.551<br>(0.077-3.940) | 1.837<br>(-0.335-4.010) | - |
| Vaccination status | No | R | R | R | R | R | R | R | R |
|  | Yes | 0.582<br>(0.196-1.726) | - | 1.044<br>(0.426-2.555) | - | 0.351<br>(0.047-2.631) | - | 0.764<br>(-2.939-4.466) | - |
| Tocilizumab | - | 1.584<br>(0.930-2.698) | - | 2.479 | 1.675 | 1.483<br>(0.725-3.032) | - | 2.815<br>(0.553-5.078) * | 2.689<br>(-0.757-6.134) |
| Variable | Response | COR (95% CI) | AOR (95% CI) | COR (95% CI) | AOR (95% CI) | COR (95% CI) | AOR (95% CI) | CB (95% CI) | AB (95% CI) |
|  |  |  |  | (1.494-4.114)<br>* | (0.294-9.524) |  |  |  |  |
| Remdisvir | - | 0.916<br>(0.621-1.352) | - | 0.591<br>(0.414-0.845)<br>* | 0.192<br>(0.044-0.835)<br>* | 1.094<br>(0.645-1.856) | - | 1.564<br>(0.043-3.085)<br>* | 0.054<br>(-2.380-2.488) |

\*P-value<0.05, R: Reference Group, COR: Crude Odds Ratio, AOR: Adjusted Odds Ratio, CB: Crude B Coefficient, AB: Adjusted B Coefficient.

### Comparison between the First and the Second Waves of the Pandemic

The analysis that compared between the first and the second waves in COVID-19 outcomes showed that there was no significant difference between the waves in regards to COVID-19 related mortality, intubation or length of hospital stay. Regarding COVID-19 related mortality, 17.7% of the patients died in the first wave while 25.8% of the patients died in the second wave. However, the second wave had significantly higher rates of ICU admission compared to the first one (Table 4; AOR=4.728; 95% CI: 3.079-7.259) with 11.3% ICU admission in the first wave and 37.6% ICU admission in the second wave. Furthermore, after adjusting for patients’ demographics and comorbidities, the second wave also had higher rates of ICU admissions compared to the first wave (Table 4; AOR=5.466; 95% CI: 3.402-8.781). In addition, the model that was adjusted for patients’ demographics, comorbidities, steroids use in the treatment and vaccination status also showed that the second wave had higher rates of ICU admissions compared to the first wave (Table 4; AOR=4.797; 95% CI: 2.903-7.927).

**Table 4.**
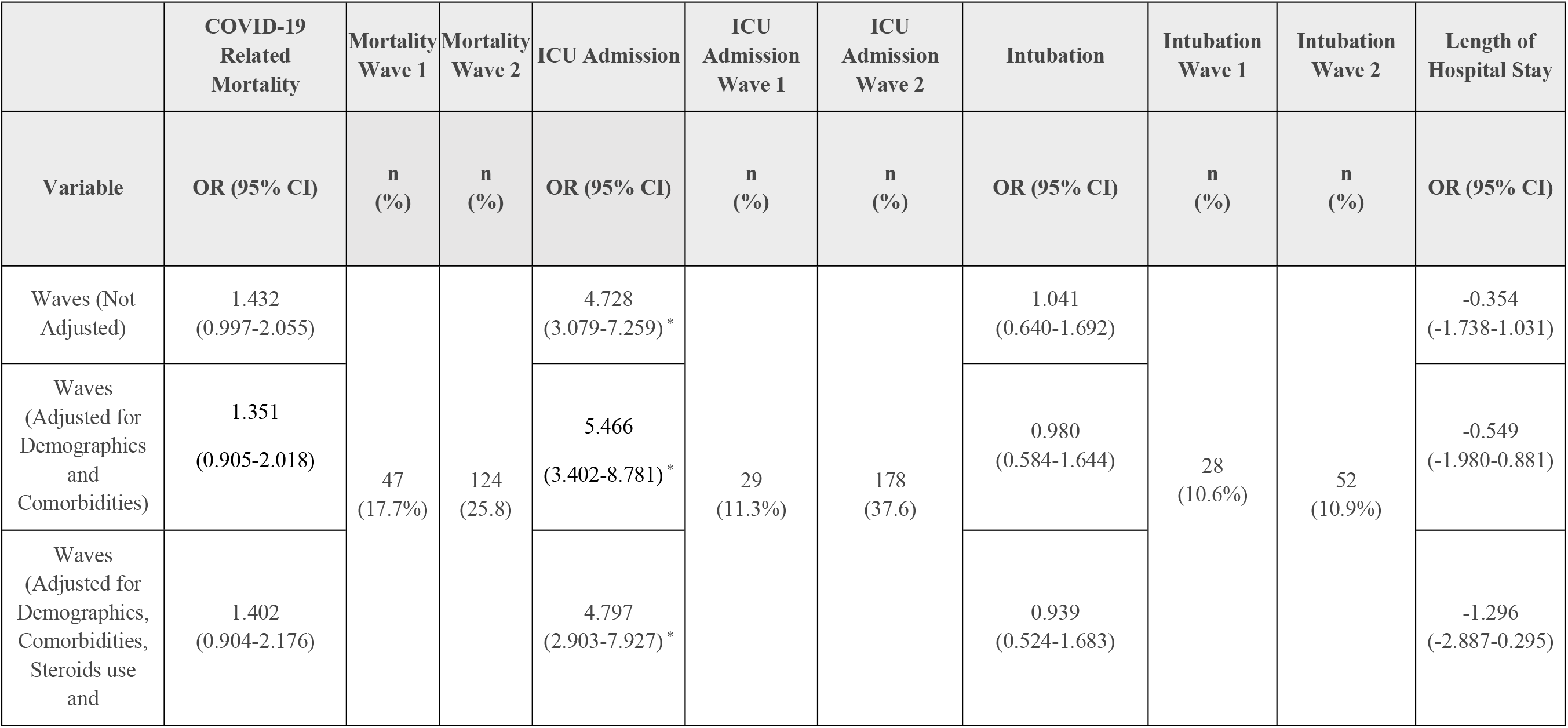
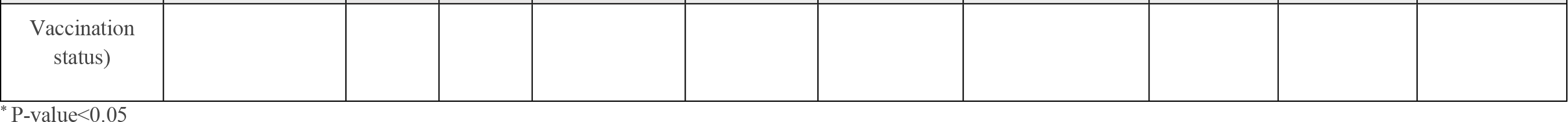
Regression Analysis for the Comparison between the First and the Second Wave of the Pandemic. Add Percentages.

## Discussion

According to our knowledge, this is the first study conducted in Jordan and one of few studies conducted in the Middle Eastern region that characterized the clinical features of COVID-19 patients and identified the factors related to COVID-19 severity and mortality.

Our study showed that the most frequent symptoms COVID-19 patients presented with were shortness of breath (35.0%), cough (15.5%) and generalized fever (15.0%). The prevalence of the COVID-19 symptoms varied between the studies as several studies in Brazil [20], Saudi Arabia [21], Kuwait [16] and China [2] reported higher prevalence of fever and cough with fever being the most common symptom among their patients. In addition, we reported a low percentage of asymptomatic patients (<1%) whereas studies in the USA [22], Kuwait [16] and China [23] reported that around 40% of their patients were asymptomatic. However, these differences can be explained by the fact that the majority of these studies were done at the beginning of the pandemic when most of the COVID-19 patients regardless of their symptom’s severity were admitted to the hospitals. Furthermore, our findings demonstrated that the most common radiological finding in chest X-rays among COVID-19 patients was bilateral infiltrates (70.9%). Similarly, studies conducted in China [2] and the United Kingdom [24] also reported that the most common finding in chest x-rays among COVID-19 patients was bilateral infiltration. In addition, most of the patients in our study who underwent HRCT imaging had ground glass opacification (70.0%) which is also consistent with previous studies conducted in China [2]. Moreover, more than 50% of the patients in our cohort required non-invasive oxygen support systems with the face masks and non-rebreather masks being the most commonly used. Whereas only 10.8% of the patients in our study needed invasive intubation. The rates of using non-invasive oxygen support systems are similar to what is reported in previous studies [24]. However, the rates of intubation in our cohort were lower compared to previous studies which reported that around 20% of the admitted patients required invasive intubation [8]. This low rate of intubation might be a consequence of refusal of intubation by the patients or the patients’ relatives due to insufficient knowledge regarding COVID-19 among the Jordanian population [25]. Moreover, mortality rates in our cohort were 23% which is quite similar to the rates reported in previous studies conducted in Kuwait [16], Spain [26] and Germany [27]. In addition, the ICU mortality rates in our study (53.1%) are consistent with several studies conducted in Italy [28], Germany [27], Saudi Arabia [21] and Kuwait [16]. However, our study had high rates of comorbidities as almost half of our study cohort had diabetes and hypertension.

Regarding the predictors of COVID-19 related mortality and severity, CKD, positive troponin, LDH and O2 saturation below 90% were significantly associated with higher risk for COVID-19 mortality. Previous studies showed that positive troponin was a significant predictor of COVID-19 mortality indicating that cardiovascular injury was associated with higher mortality among COVID-19 patients [29]. However, in contrast to our study, studies showed that another cardiac marker, BNP was also associated with higher risk for mortality [30]. Additionally, LDH was significantly associated with higher risk of mortality which was consistent with previous studies that showed that higher LDH levels were independent predictors for mortality [29]. LDH is an enzyme that is found in many organs with its elevation indicating multiorgan injuries [31]. Although in our study diabetes and hypertension were not predictors of mortality which contradict the results of several studies [16], our study showed that CKD, which is a common complication of hypertension and diabetes was a major predictor of COVID-19 mortality. Similar to our study, several articles showed that low O2 saturation at presentation was associated with higher COVID-19 mortality as low O2 saturation is associated with higher degree of lung injury and more delayed presentation [32]. Also, our analysis showed that low O2 saturation was a significant predictor of invasive intubation. Surprisingly, the results revealed that the presentation with GI symptoms was associated with lower risk of invasive intubation. A meta-analysis showed that the prevalence of GI symptoms among COVID-19 patients was 20% and it was not associated with higher risk for mortality [33]. In addition, a cohort study showed that patients who presented with GI symptoms had a significantly lower COVID-19 severity [34]. Furthermore, our analysis revealed that heart failure and the use of remdesivir in treating patients were the only significant factors associated with ICU admission. Studies showed that 1 in 4 patients with heart failure hospitalized for COVID-19 infection died [35]. Moreover, consistent with our findings, cohort studies showed that remdesivir reduced COVID-19 ICU admission but not mortality [36, 37].

It is important to mention that in our study age and sex were not significantly associated with higher COVID-19 severity or mortality. The majority of the studies in the literature showed that older age is a significant predictor of COVID-19 severity and mortality [38]. However, our findings can be explained by the fact that half of our patients were above 65 with a mean age of 63.15 for the whole cohort which limits the comparison in COVID-19 outcomes between elderly and young patients. On the other hand, the results about the association between COVID-19 mortality and sex were inconsistent in the literature with some studies reporting a higher risk of mortality among male sex [39] and others reporting a lack of a significant difference between males and females in the rates of COVID-19 mortality [40]. Additionally, our study showed that vaccination status was not a significant predictor of COVID-19 mortality or severity which might be explained by the fact that at the time of conducting this study COVID-19 vaccines were not widely available as only 3.2% of our study cohort were vaccinated. Despite the fact that several studies showed that steroids (dexamethasone and methylprednisolone) use among COVID-19 patients reduced COVID-19 severity and mortality [41], our study showed no such association. Yet, in our study, steroids were used to treat a very high percentage of patients (88.6%) which limits the comparison in the outcomes between patients who were and were not treated with steroids.

Our study showed that the second wave of the pandemic was associated with higher rate of ICU admissions but not intubation or mortality compared to the first wave. Even after adjusting for patients’ characteristics, vaccination status and methods of treatment, the second wave had significantly higher rates of ICU admissions. These findings can be explained by the fact that the second wave was driven by the transmission of the delta and alpha variants which might indicate that these variants were associated with higher disease severity compared to the original variant that spread in Wuhan. Studies showed that the Alpha variant was significantly associated with higher rates of mortality in multiple European countries [42, 43]. However, none of them found that it was also associated with higher ICU admission rates.

However, this study has several limitations. First, this was a single center cohort study which limits the generalizability of its findings. Second, although our analysis was adjusted for confounding variables, the risk of confounding bias cannot be totally excluded. Third, we did not perform genetic sequencing to prove the SARS-COV-2 variants that affected the patients during the two waves of the pandemic which might limit the interpretations of our analysis that compared the 2 waves disease severity and mortality.

## Conclusion

To conclude, we provide the detailed clinical characteristics, investigations and treatment methods of 745 COVID-19 patients. The overall rates of COVID-19 related mortality, ICU admission and invasive intubation were 23.0%, 28.3% and 10.8%, respectively. Our study showed that CKD, positive troponin, LDH and O2 saturation<90% at admission were significant predictors of COVID-19 morality. Moreover, heart failure patients had a significantly higher risk for ICU admission while remdesivir use in treating COVID-19 patients was associated with a reduction in the risk of ICU admission. Patients who had O2 saturation<90% at admission had a significantly higher risk for invasive intubation whereas patients presenting with GI symptoms had a significantly lower risk for it. Additionally, the second wave of the pandemic had significantly higher rates of ICU admission but not mortality or invasive intubation compared to the first wave.

## Data Availability

The data associated with this manuscript are available in the Supplementary Material

## Contributions

KA and ASA were involved in Conceptualization; KA, ASA, AAT, MKR, MMH, AHA, DLA, MR and NO were involved in Data curation, Formal analysis, Investigation, Methodology, Project administration, Resources, Software, Validation, Visualization, and Writing the original draft; ASA, KA, AAT and NO were involved in Supervision and Reviewing & Editing the manuscript.

## Data Sharing

The data associated with this manuscript are available in the Supplementary Material.

## Acknowledgments

None.

